# MicroRNAs as diagnostic biomarkers and predictors of antidepressant response in major depressive disorder: a systematic review

**DOI:** 10.1101/2024.02.17.24302977

**Authors:** Beatriz A. Carneiro, Lívia N. F. Guerreiro-Costa, Daniel H. Lins-Silva, Daniela Faria-Guimarães, Lucca S. Souza, Gustavo C. Leal, Ana Teresa C. Fontes, Graziele Beanes, Ryan S. Costa, Lucas C. Quarantini

## Abstract

Despite the hardships of major depressive disorder (MDD), biomarkers for the diagnosis and pharmacological management of this condition are lacking. MicroRNAs are epigenetic mechanisms that could provide promising MDD biomarkers. Our aim was to summarize the findings and provide validation for the selection and use of specific microRNAs as biomarkers in the diagnosis and treatment of MDD. A systematic review was conducted using the PubMed/MEDLINE, Cochrane, PsycINFO, Embase, and LILACS databases from March to May 2022, with clusters of terms based on “microRNA” and “antidepressant”. Studies involving human subjects, animal models, and cell cultures were included, whereas those that evaluated herbal medicines, non-pharmacological therapies, or epigenetic mechanisms other than miRNA were excluded. The review revealed differences in the expression of various microRNAs when considering the time of assessment (before or after antidepressant treatment) and the population studied. However, due to the heterogeneity of the microRNAs investigated, the limited size of the samples, and the wide variety of antidepressants used, few conclusions could be made. Despite the observed heterogeneity, the following microRNAs were determined to be important factors in MDD and the antidepressant response: mir-1202, mir-135, mir-124, and mir-16. The findings indicate the potential for the use of microRNAs as biomarkers for the diagnosis and treatment of MDD; however, more homogeneous studies are needed.

## 1. Introduction

Major depressive disorder (MDD) is a debilitating disorder that has a prevalence rate of greater than 10% [1-2] and a remission rate after conventional treatment of approximately 50% [3]. Genetic variants are involved in the pathophysiology of mental disorders and possibly in the treatment response, with a study reporting that 42% of the variation in outcomes after treatment can be attributed to genetic factors [4]. Therefore, a greater understanding of the genetics involved in MDD can help optimize pharmacological treatments by increasing the accuracy of drug response and tolerability predictions.

Our current understanding of the molecular pathophysiology of MDD is limited, although studies have associated MDD with genetic polymorphisms, epigenetic mechanisms, and synaptic plasticity alterations [5-7]. Some genetic features are inherited; however, most are modified during prenatal development and throughout the lifetime by various environmental factors, including food and drug exposure [8]. Three epigenetic mechanisms function in the modulation of gene expression [9]. Two of these change the DNA structure without affecting its sequence (DNA methylation and histone acetylation) and the third modulates gene expression at the post-transcriptional level (miRNAs) [9].

MicroRNAs (miRNAs) are small non-coding RNAs that influence and regulate gene expression, thereby acting as epigenetic modulators [10]. By binding to specific genes, these miRNAs affect neural plasticity and brain function by inhibiting translation or degrading mRNA [10]. Previous animal studies found that stress, which is a risk factor for MDD, interfered with miRNA expression in rodent brains. This finding supports the involvement of miRNAs in the etiopathogenesis of MDD, highlighting a possible role for these molecules in the diagnosis of psychiatric diseases. Furthermore, other studies have identified strong associations between certain miRNAs and antidepressant (AD) responses, including miR-1202, miR-124, miR-135a, miR-145, and miR-20b, which are promising response biomarker candidates [11]. Lopez et al. [12] demonstrated that miR-146a-5p, miR-146b-5p, miR-425-3p, and miR-24-3p levels decreased following AD treatment. However, the expression of miRNAs between different body components and the relationship between the peripheral and central levels of this biomolecule are inconsistent.

The pathophysiology of MDD is a promising ongoing topic of research. However, we believe that the contribution of miRNAs is under-explored, despite the considerable interest in obtaining biological biomarkers for MDD diagnosis and antidepressant treatment response [13-16]. The purpose of our review was to investigate changes in miRNA expression before and after pharmacological treatment for MDD.

## 2. Methods

We conducted a systematic review of the literature following the recommendations of the Cochrane handbook [17] and preferred reporting items for systematic reviews and meta-analyses (PRISMA) guidelines [18]. The review was registered in PROSPERO (CRD42021265268).

### 2.1. Literature search and study selection

From March 2022 through March 2023, we searched the PubMed/MEDLINE, Cochrane, PsycINFO, Embase, and LILACS databases using the following terms: (micrornas OR microrna OR mirnas OR micro rna) AND (antidepressive agents OR antidepressive drugs OR antidepressive agent OR antidepressive OR antidepressants OR antidepressant OR antidepressant drugs OR antidepressant OR antidepressant drug OR antidepressive treatment). No limits to publication years were added. The articles were required to be published in English and to involve humans, animals, or cell cultures. The titles and abstracts of the retrieved items were independently screened by four reviewers (LG, BC, DH, LS), and the two main reviewers (LG, BC) then independently assessed the full texts of the selected studies.

### 2.2. Eligibility criteria

The strategic question that guided the manual search and the inclusion and exclusion criteria was as follows: “What is the levels of miRNAs expression before and after AD treatment?” The inclusion criteria were (1) original studies only; (2) human subjects aged 18 or over with a clinical diagnosis of MDD and undergoing treatment with ADs; and (3) animal models and cell cultures with miRNA assessments after treatment with ADs. For each study, we prepared a standardized spreadsheet that presented the following information: i) authors and publication year, ii) publication journal, iii) population studied, iv) treatment conducted, v) instrument used for diagnosis and stratification of MDD, vi) study design, vii) miRNA extraction site, viii) follow-up time, and ix) miRNA expression before and after intervention. We excluded studies that evaluated herbal medicines, non-pharmacological therapies, or epigenetic mechanisms other than miRNAs.

### 2.3. Risk of bias assessment

The quality of the study was evaluated by two reviewers (DF, DH) using the strengthening the reporting of observational studies in epidemiology (STROBE) [19] and systematic review center for laboratory animal experimentation (SYRCLE) [20] tools for the human and animal studies, respectively (Tables *1* and 2). We did not assess the quality of the cell culture studies because no standardized tool exists for this purpose. Discrepancies between the two reviewers were resolved through discussions until a consensus was reached.

**Table 1:**
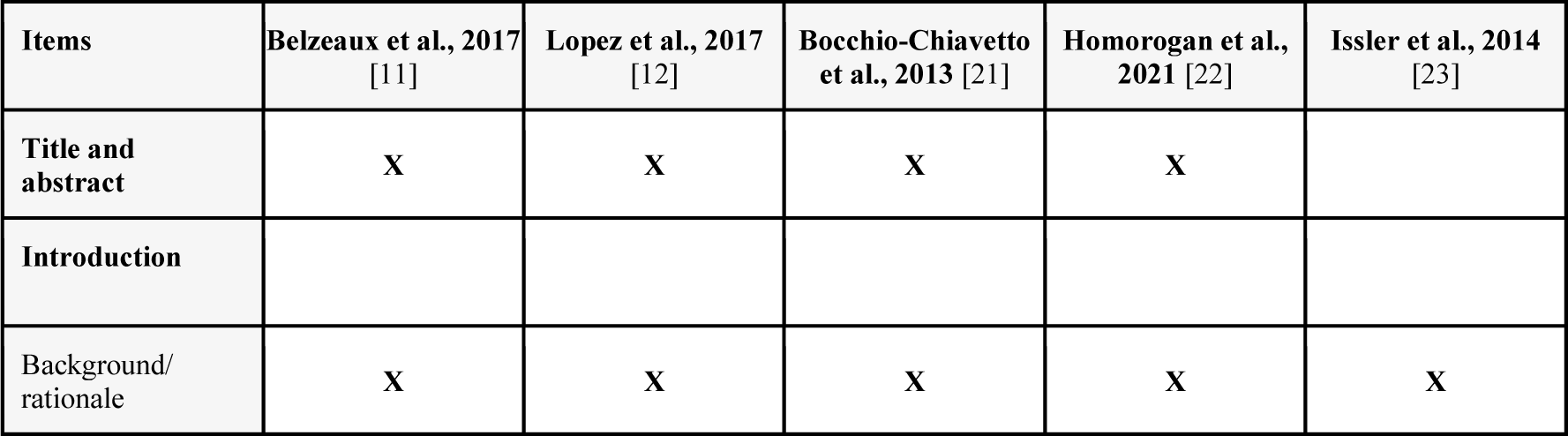
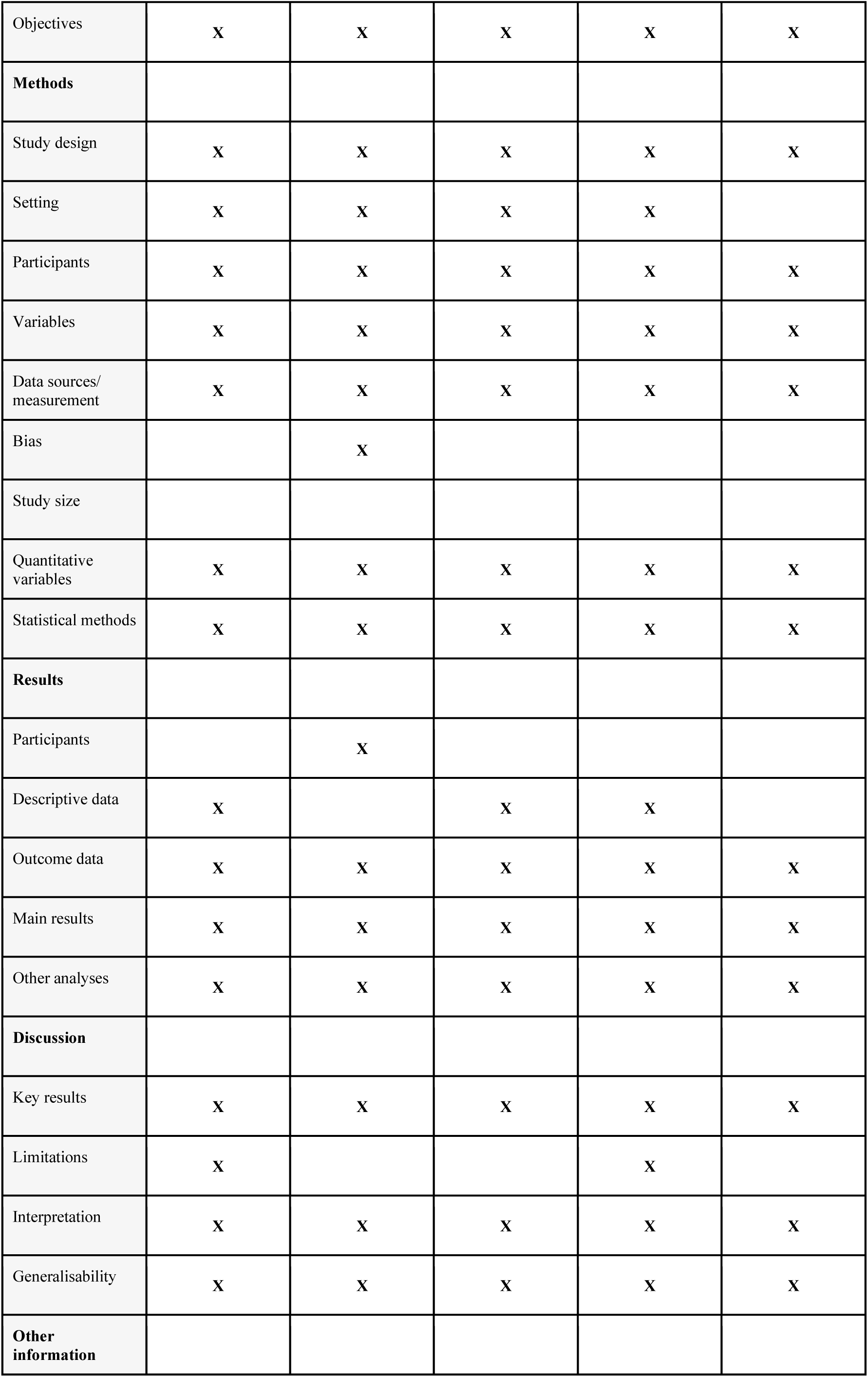
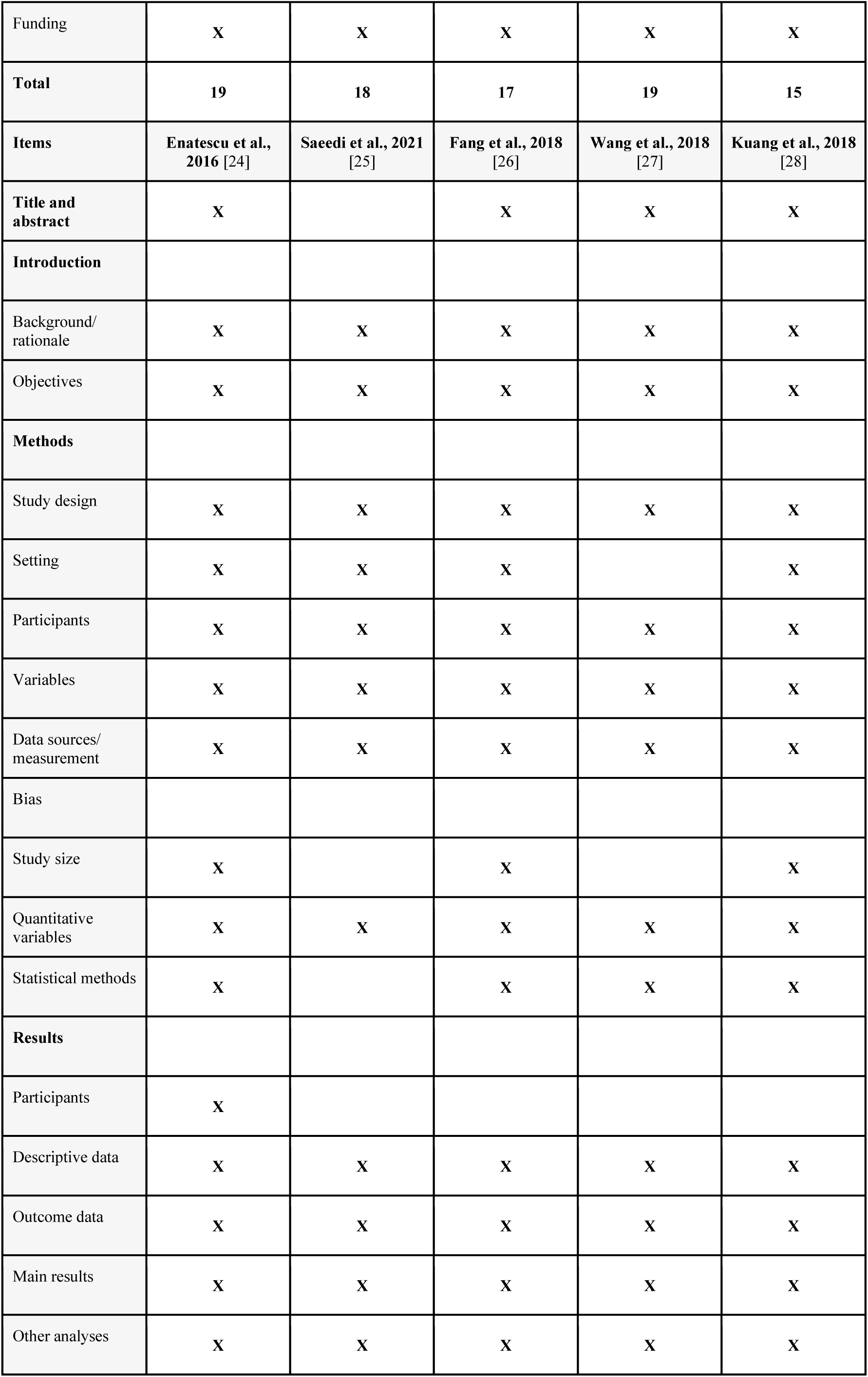
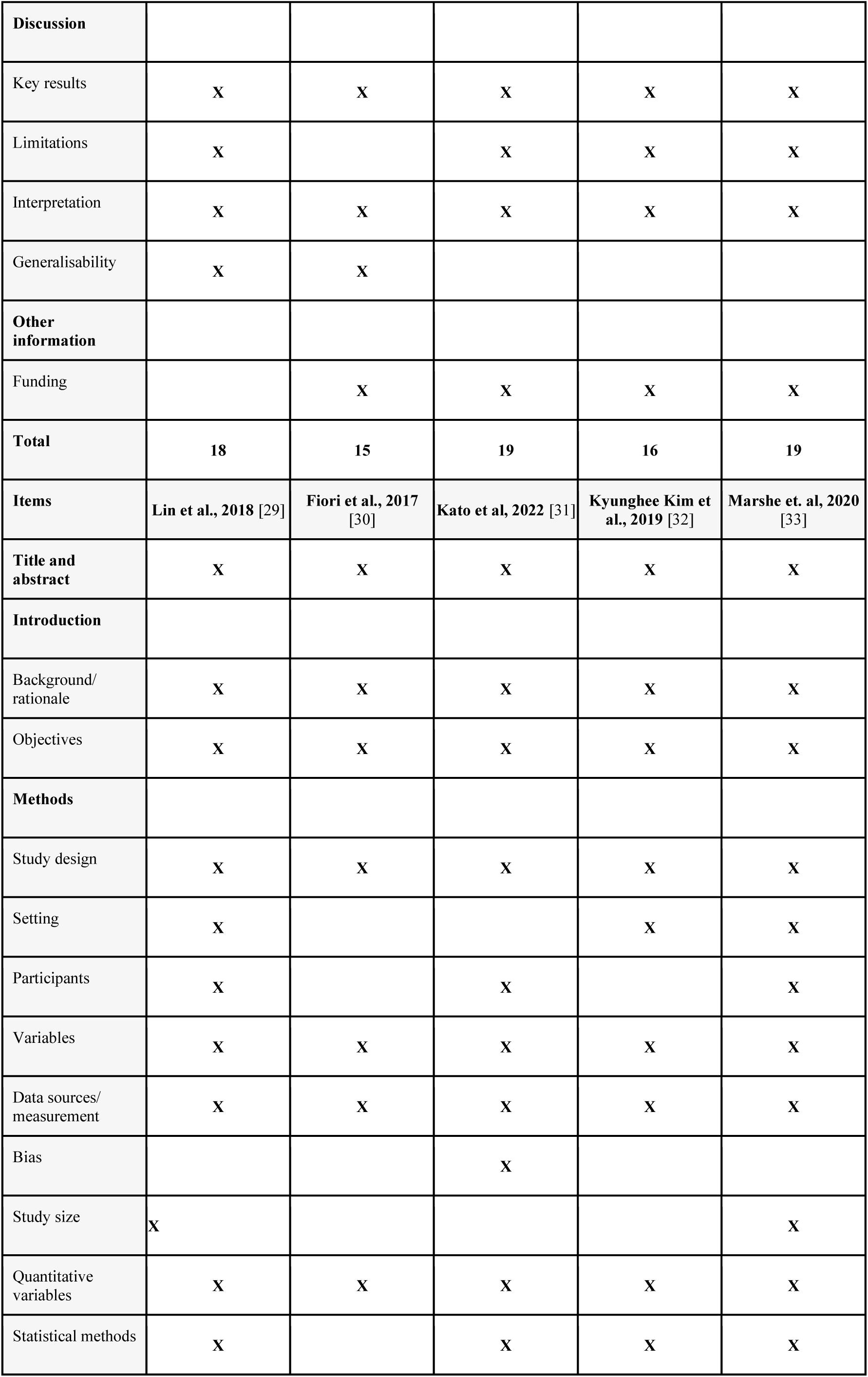
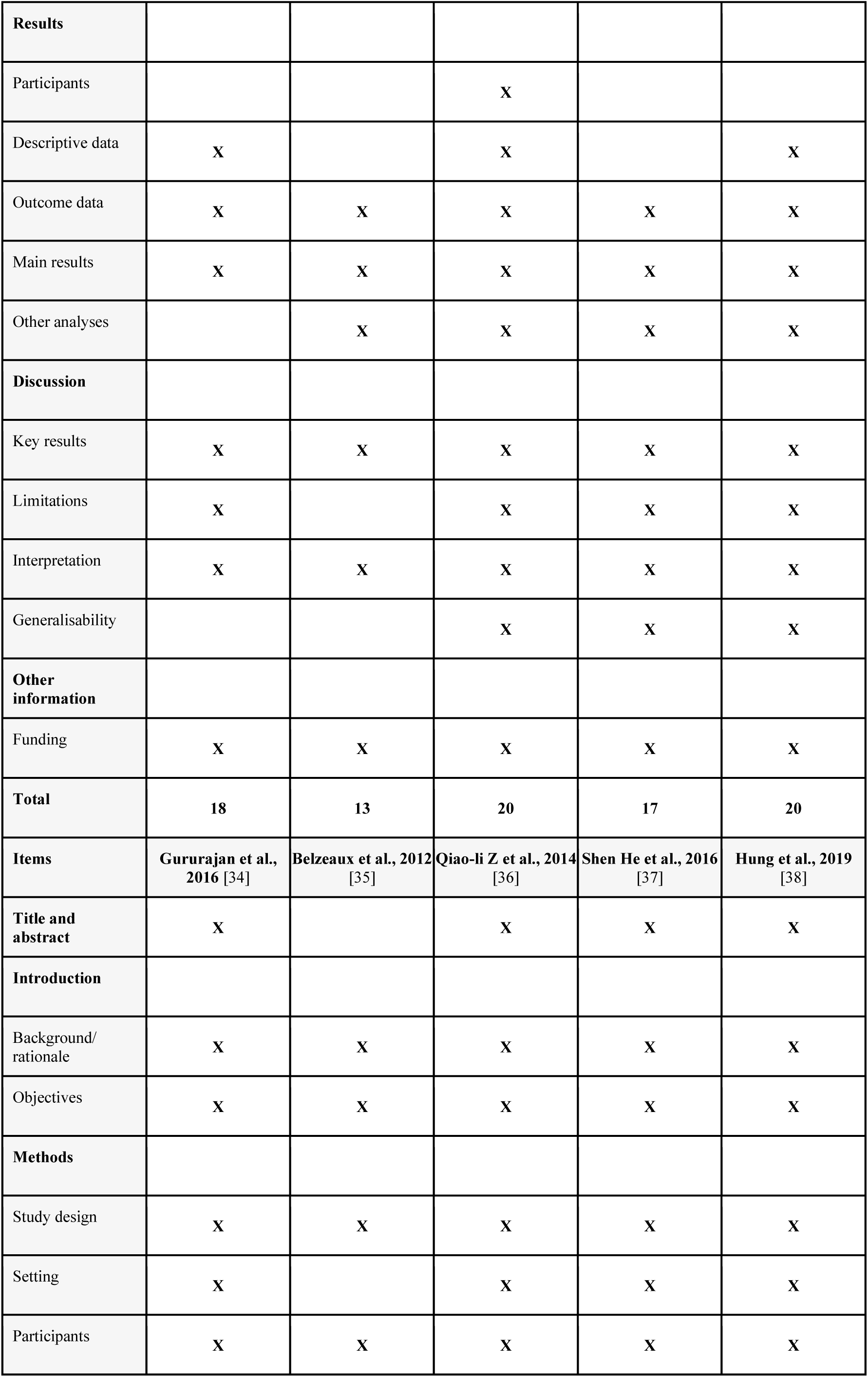
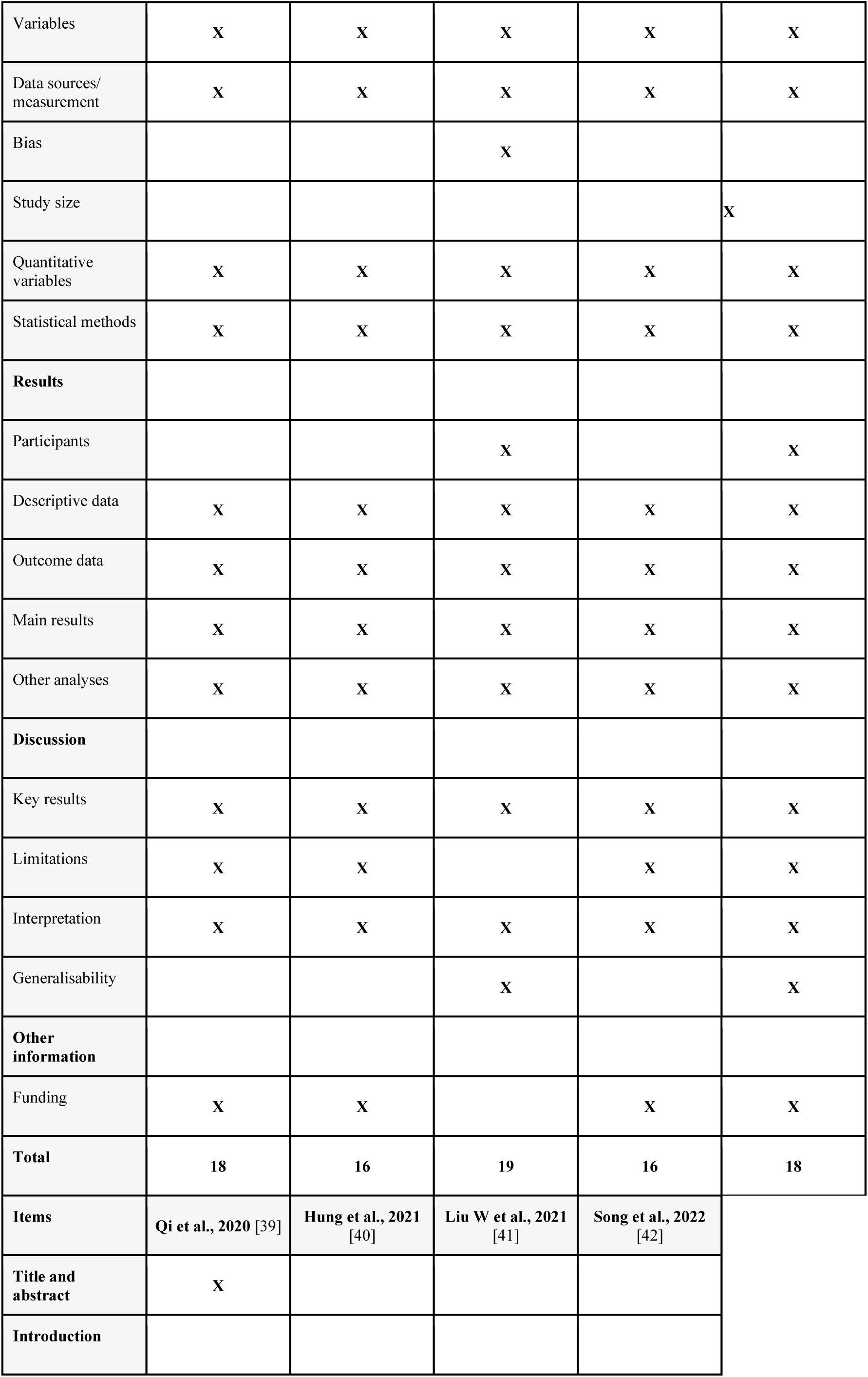
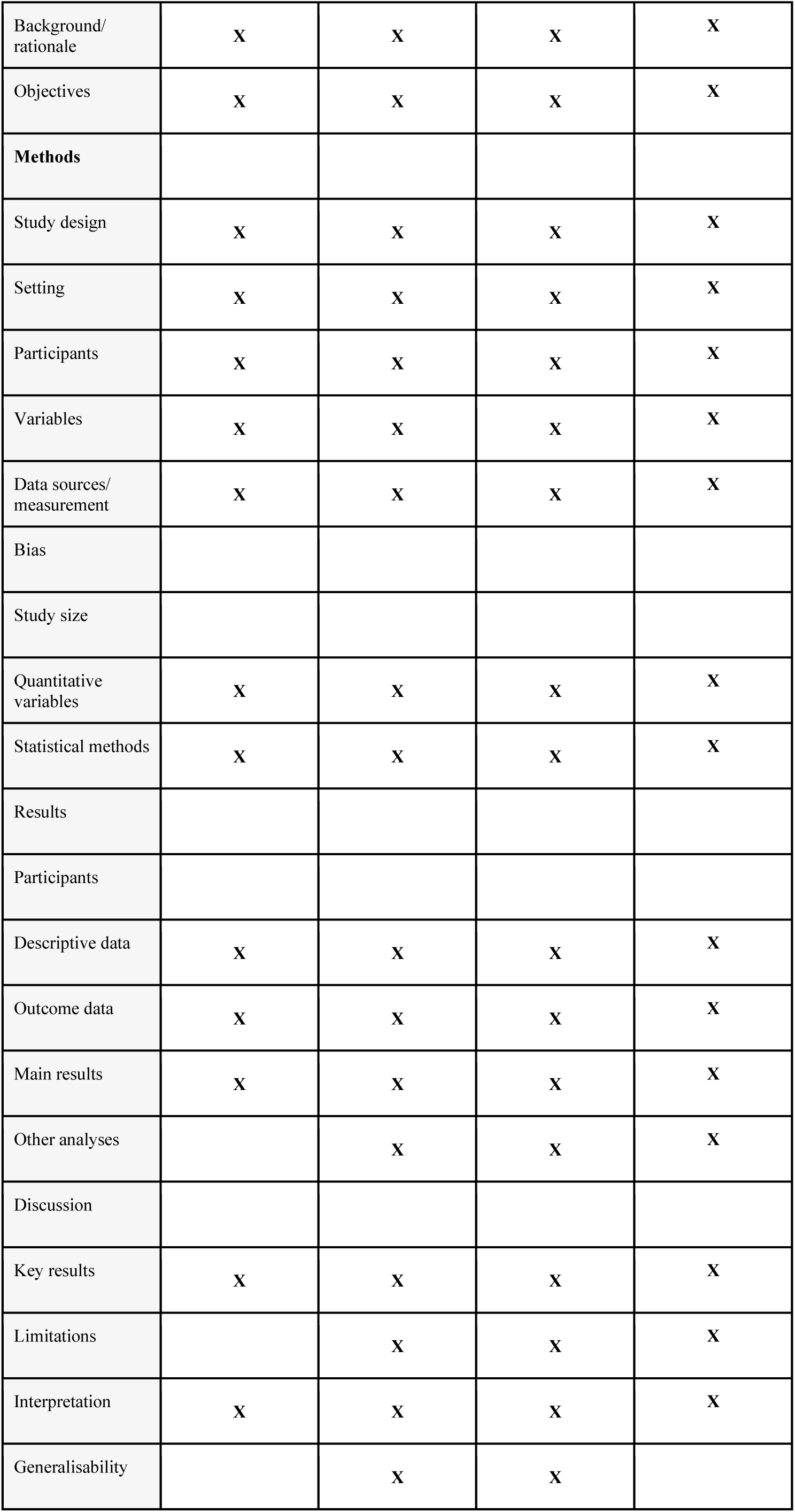
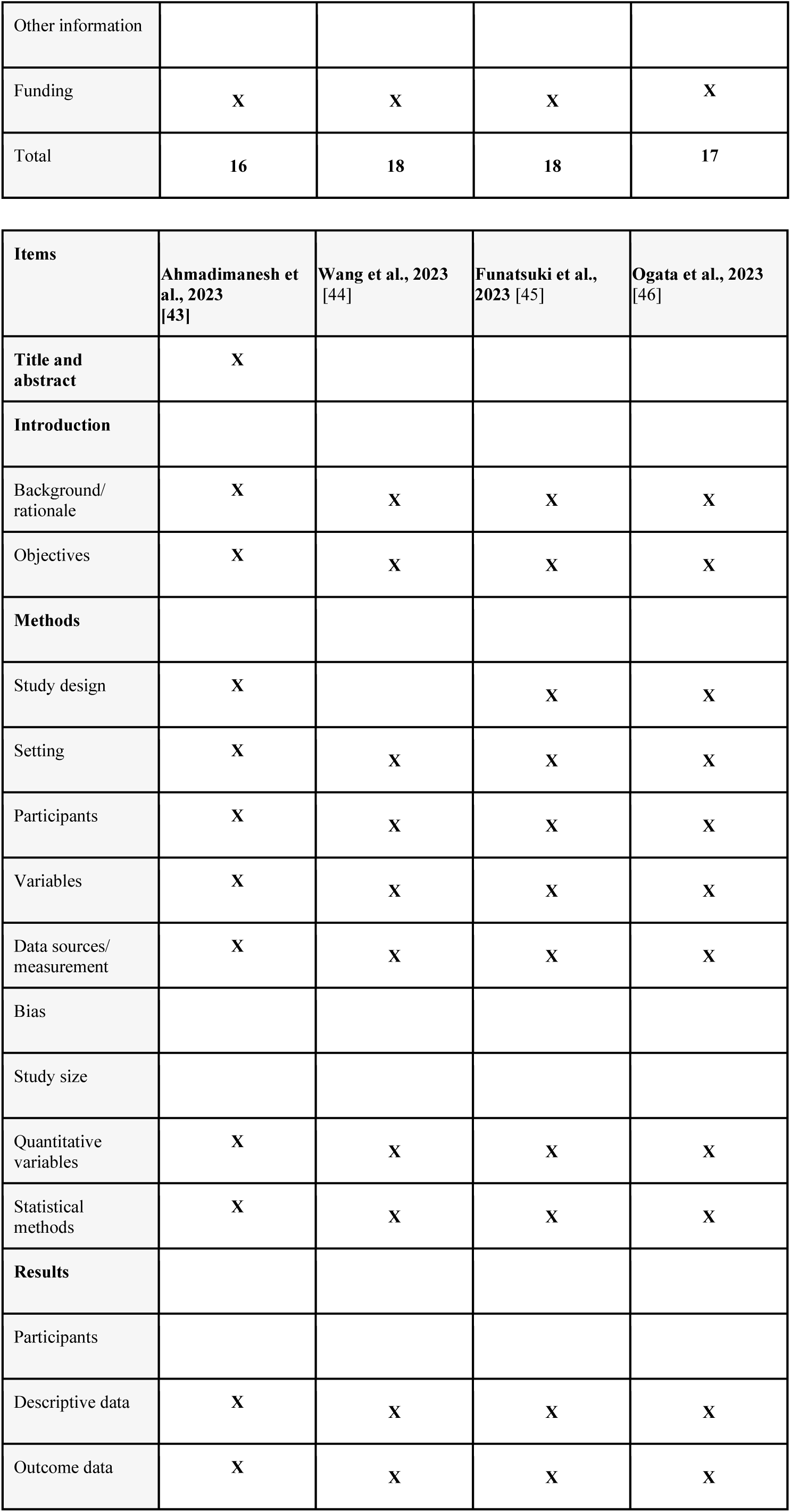
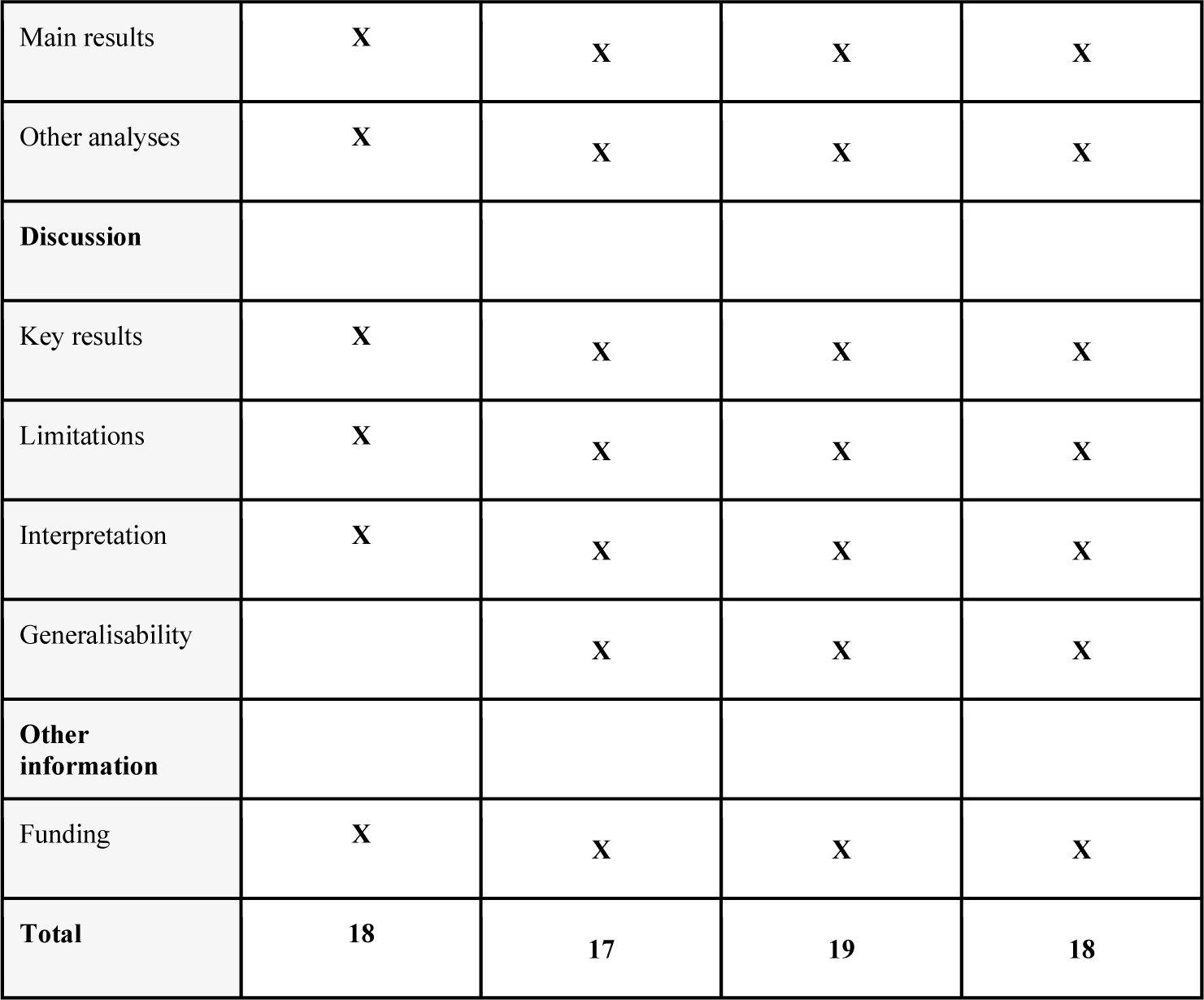
Strengthening the Reporting of Observational Studies in Epidemiology (STROBE) for human studies.

| Items | Belzeaux et al., 2017 [11] | Lopez et al., 2017 [12] | Bocchio-Chiavetto et al., 2013 [21] | Homorogan et al., 2021 [22] | Issler et al., 2014 [23] |
| --- | --- | --- | --- | --- | --- |
| Title and abstract | X | X | X | X |  |
| Introduction |  |  |  |  |  |
| Background/rationale | X | X | X | X | X |
| Objectives | X | X | X | X | X |
| **Methods** |  |  |  |  |  |
| Study design | X | X | X | X | X |
| Setting | X | X | X | X |  |
| Participants | X | X | X | X | X |
| Variables | X | X | X | X | X |
| Data sources/<br>measurement | X | X | X | X | X |
| Bias |  | X |  |  |  |
| Study size |  |  |  |  |  |
| Quantitative<br>variables | X | X | X | X | X |
| Statistical methods | X | X | X | X | X |
| **Results** |  |  |  |  |  |
| Participants |  | X |  |  |  |
| Descriptive data | X |  | X | X |  |
| Outcome data | X | X | X | X | X |
| Main results | X | X | X | X | X |
| Other analyses | X | X | X | X | X |
| **Discussion** |  |  |  |  |  |
| Key results | X | X | X | X | X |
| Limitations | X |  |  | X |  |
| Interpretation | X | X | X | X | X |
| Generalisability | X | X | X | X | X |
| **Other**<br>**information** |  |  |  |  |  |
| Funding | X | X | X | X | X |
| **Total** | **19** | **18** | **17** | **19** | **15** |
| **Items** | **Enatescu et al.,**<br>**2016 [24]** | **Saeedi et al., 2021**<br>**[25]** | **Fang et al., 2018**<br>**[26]** | **Wang et al., 2018**<br>**[27]** | **Kuang et al., 2018**<br>**[28]** |
| **Title and abstract** | X |  | X | X | X |
| **Introduction** |  |  |  |  |  |
| Background/<br>rationale | X | X | X | X | X |
| Objectives | X | X | X | X | X |
| **Methods** |  |  |  |  |  |
| Study design | X | X | X | X | X |
| Setting | X | X | X |  | X |
| Participants | X | X | X | X | X |
| Variables | X | X | X | X | X |
| Data sources/<br>measurement | X | X | X | X | X |
| Bias |  |  |  |  |  |
| Study size | X |  | X |  | X |
| Quantitative<br>variables | X | X | X | X | X |
| Statistical methods | X |  | X | X | X |
| **Results** |  |  |  |  |  |
| Participants | X |  |  |  |  |
| Descriptive data | X | X | X | X | X |
| Outcome data | X | X | X | X | X |
| Main results | X | X | X | X | X |
| Other analyses | X | X | X | X | X |
| **Discussion** |  |  |  |  |  |
| Key results | X | X | X | X | X |
| Limitations | X |  | X | X | X |
| Interpretation | X | X | X | X | X |
| Generalisability | X | X |  |  |  |
| **Other information** |  |  |  |  |  |
| Funding |  | X | X | X | X |
| **Total** | **18** | **15** | **19** | **16** | **19** |
| **Items** | **Lin et al., 2018 [29]** | **Fiori et al., 2017 [30]** | **Kato et al, 2022 [31]** | **Kyunghee Kim et al., 2019 [32]** | **Marshe et. al, 2020 [33]** |
| **Title and abstract** | X | X | X | X | X |
| **Introduction** |  |  |  |  |  |
| Background/<br>rationale | X | X | X | X | X |
| Objectives | X | X | X | X | X |
| **Methods** |  |  |  |  |  |
| Study design | X | X | X | X | X |
| Setting | X |  |  | X | X |
| Participants | X |  | X |  | X |
| Variables | X | X | X | X | X |
| Data sources/<br>measurement | X | X | X | X | X |
| Bias |  |  | X |  |  |
| Study size | X |  |  |  | X |
| Quantitative variables | X | X | X | X | X |
| Statistical methods | X |  | X | X | X |
| **Results** |  |  |  |  |  |
| Participants |  |  | X |  |  |
| Descriptive data | X |  | X |  | X |
| Outcome data | X | X | X | X | X |
| Main results | X | X | X | X | X |
| Other analyses |  | X | X | X | X |
| **Discussion** |  |  |  |  |  |
| Key results | X | X | X | X | X |
| Limitations | X |  | X | X | X |
| Interpretation | X | X | X | X | X |
| Generalisability |  |  | X | X | X |
| **Other information** |  |  |  |  |  |
| Funding | X | X | X | X | X |
| **Total** | **18** | **13** | **20** | **17** | **20** |
| **Items** | **Gururajan et al., 2016 [34]** | **Belzeaux et al., 2012 [35]** | **Qiao-li Z et al., 2014 [36]** | **Shen He et al., 2016 [37]** | **Hung et al., 2019 [38]** |
| **Title and abstract** | X |  | X | X | X |
| **Introduction** |  |  |  |  |  |
| Background/<br>rationale | X | X | X | X | X |
| Objectives | X | X | X | X | X |
| **Methods** |  |  |  |  |  |
| Study design | X | X | X | X | X |
| Setting | X |  | X | X | X |
| Participants | X | X | X | X | X |
| Variables | X | X | X | X | X |
| Data sources/<br>measurement | X | X | X | X | X |
| Bias |  |  | X |  |  |
| Study size |  |  |  |  | X |
| Quantitative<br>variables | X | X | X | X | X |
| Statistical methods | X | X | X | X | X |
| **Results** |  |  |  |  |  |
| Participants |  |  | X |  | X |
| Descriptive data | X | X | X | X | X |
| Outcome data | X | X | X | X | X |
| Main results | X | X | X | X | X |
| Other analyses | X | X | X | X | X |
| **Discussion** |  |  |  |  |  |
| Key results | X | X | X | X | X |
| Limitations | X | X |  | X | X |
| Interpretation | X | X | X | X | X |
| Generalisability |  |  | X |  | X |
| **Other**<br>**information** |  |  |  |  |  |
| Funding | X | X |  | X | X |
| **Total** | **18** | **16** | **19** | **16** | **18** |
| **Items** | Qi et al., 2020 [39] | Hung et al., 2021<br>[40] | Liu W et al., 2021<br>[41] | Song et al., 2022<br>[42] |  |
| **Title and**<br>**abstract** | X |  |  |  |  |
| **Introduction** |  |  |  |  |  |

|  |  |  |  |  |
| --- | --- | --- | --- | --- |
| Background/<br>rationale | X | X | X | X |
| Objectives | X | X | X | X |
| **Methods** |  |  |  |  |
| Study design | X | X | X | X |
| Setting | X | X | X | X |
| Participants | X | X | X | X |
| Variables | X | X | X | X |
| Data sources/<br>measurement | X | X | X | X |
| Bias |  |  |  |  |
| Study size |  |  |  |  |
| Quantitative<br>variables | X | X | X | X |
| Statistical methods | X | X | X | X |
| Results |  |  |  |  |
| Participants |  |  |  |  |
| Descriptive data | X | X | X | X |
| Outcome data | X | X | X | X |
| Main results | X | X | X | X |
| Other analyses |  | X | X | X |
| Discussion |  |  |  |  |
| Key results | X | X | X | X |
| Limitations |  | X | X | X |
| Interpretation | X | X | X | X |
| Generalisability |  | X | X |  |
| Other information |  |  |  |  |
| Funding | X | X | X | X |
| Total | 16 | 18 | 18 | 17 |

| Items | Ahmadimanesh et al., 2023 [43] | Wang et al., 2023 [44] | Funatsuki et al., 2023 [45] | Ogata et al., 2023 [46] |
| --- | --- | --- | --- | --- |
| Title and abstract | X |  |  |  |
| Introduction |  |  |  |  |
| Background/rationale | X | X | X | X |
| Objectives | X | X | X | X |
| Methods |  |  |  |  |
| Study design | X |  | X | X |
| Setting | X | X | X | X |
| Participants | X | X | X | X |
| Variables | X | X | X | X |
| Data sources/measurement | X | X | X | X |
| Bias |  |  |  |  |
| Study size |  |  |  |  |
| Quantitative variables | X | X | X | X |
| Statistical methods | X | X | X | X |
| Results |  |  |  |  |
| Participants |  |  |  |  |
| Descriptive data | X | X | X | X |
| Outcome data | X | X | X | X |
| Main results | X | X | X | X |
| Other analyses | X | X | X | X |
| **Discussion** |  |  |  |  |
| Key results | X | X | X | X |
| Limitations | X | X | X | X |
| Interpretation | X | X | X | X |
| Generalisability |  | X | X | X |
| **Other information** |  |  |  |  |
| Funding | X | X | X | X |
| **Total** | **18** | **17** | **19** | **18** |

**Table 2:** Systematic Review Center for Laboratory animal Experimentation (SYRCLE) for animal studies.

| Type of bias | Domain | Issler et al., 2014 [23] | Patrício et al, 2020 [47] | Lo Iacono et al., 2021 [48] | Zhang et al., 2015 [49] | Song et al., 2019 [50] | Xie et al., 2019 [51] | Grieco et al., 2017 [52] |
| --- | --- | --- | --- | --- | --- | --- | --- | --- |
| Selection bias | Sequence generation | No | Unclear | Unclear | Unclear | Yes | No | No |
|  | *Baseline characteristics* | *Yes* | *Yes* | Yes | Yes | *No* | *Yes* | Yes |
|  | *Allocation concealment* | *Unclear* | *Unclear* | Unclear | Unclear | *Unclear* | *Unclear* | Unclear |
| Performance bias | *Random housing* | *Yes* | *Yes* | Yes | Yes | *Unclear* | *Yes* | Yes |
|  | *Blinding* | *Unclear* | *Yes* | Unclear | Yes | *Unclear* | *Unclear* | Unclear |
| Detection bias | *Random outcome assessment* | *Unclear* | *Unclear* | Unclear | Yes | *Unclear* | *Unclear* | Unclear |
|  | *Blinding* | *Yes* | *Yes* | Yes | Yes | *Unclear* | *Yes* | Yes |
| Attrition bias | *Incomplete outcome data* | *Unclear* | *Unclear* | Unclear | Unclear | *Yes* | *No* | Unclear |
| Reporting bias | *Selective outcome reporting* | *Unclear* | *Yes* | Yes | Yes | *Yes* | *Unclear* | Yes |
| Other | *Other sources of bias* | *Yes* | *Yes* | Unclear | Yes | *Yes* | *Unclear* | Yes |

| Type of bias | Domain | Wan et al., 2018 [53] | Mingardi, 2021 [54] | Huang et al., 2021 [55] | Pan et al., 2015 [56] | Higuchi et al., 2016 [57] | Zheng et al., 2023 [58] | Guan et al., 2023 [59] |
| --- | --- | --- | --- | --- | --- | --- | --- | --- |
| Selection bias | Sequence generation | Unclear | Unclear | No | Yes | No | Unclear | Unclear |
|  | Baseline characteristics | Yes | Yes | Yes | No | Yes | Yes | Yes |
|  | Allocation concealment | Unclear | Unclear | Unclear | Unclear | Unclear | Unclear | Unclear |
| Performance bias | Random housing | No | Yes | Yes | Unclear | Unclear | Yes | Unclear |
|  | Blinding | No | Yes | Unclear | Unclear | Unclear | Unclear | Unclear |
| Detection bias | Random outcome assessment | Unclear | Yes | Unclear | Unclear | Unclear | Unclear | Unclear |
|  | Blinding | Yes | Yes | Unclear | Yes | Yes | Unclear | Unclear |
| Attrition bias | Incomplete outcome data | Unclear | No | No | Yes | Unclear | Unclear | Unclear |
| Reporting bias | Selective outcome reporting | Yes | Yes | Yes | Yes | Yes | Yes | Unclear |
| Other | Other sources of bias | Yes | Yes | Yes | Yes | Yes | Yes | Yes |

## 3. Results

The initial search of the databases returned 6,246 articles, and after reviewing the titles, 201 of these were selected for abstract perusal. A full reading was performed on the 84 studies selected based on the abstract, and 42 of these were included in the analysis and data extraction procedure (Figure *1*): 15 studies conducted in China; 7 in Canada; 4 in Japan; 3 each in Italy, Taiwan, and Israel; 2 in Romania; and 1 each in Portugal, Ireland, France, Iran and the United States. The majority were human studies that used data from cohorts or clinical trials.

### 3.1. Human beings

We found 28 studies comparing the miRNA expressions of people undergoing AD treatment (Table *3*). The miRNA levels of patients were assessed following various therapeutic regimens. In eleven studies, participants were treated exclusively with selective serotonin reuptake inhibitors (SSRIs), with escitalopram/citalopram drugs used in eight of these.

**Table 3.**
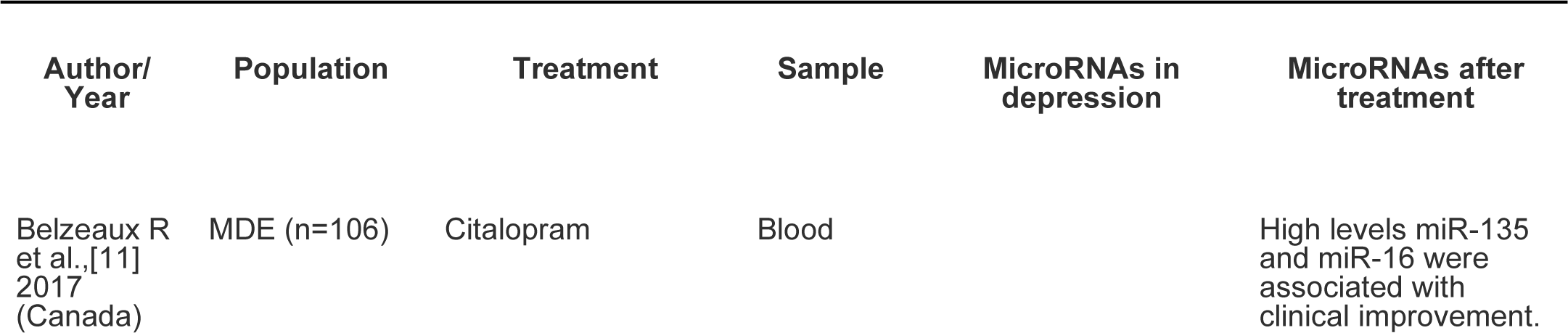
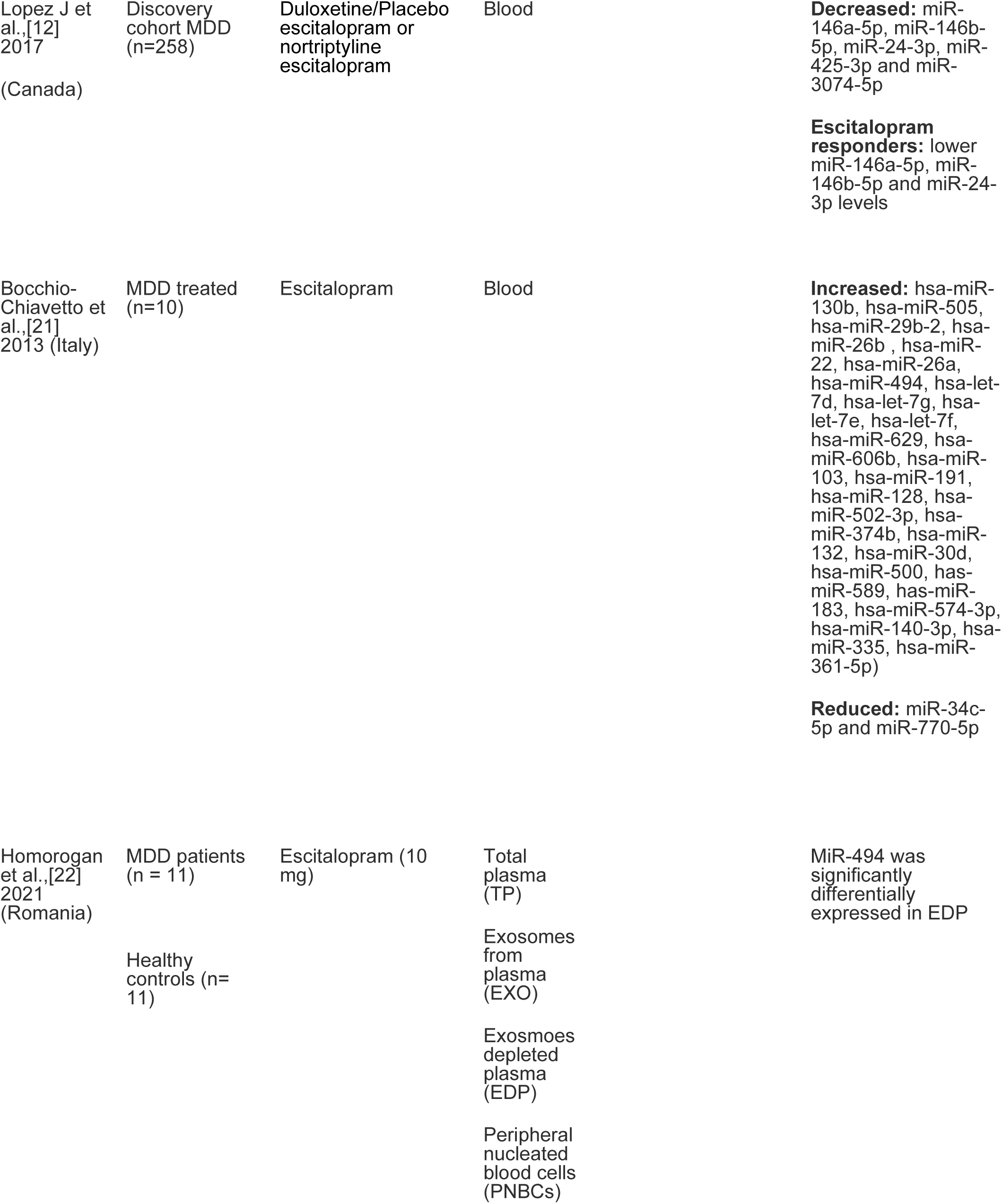
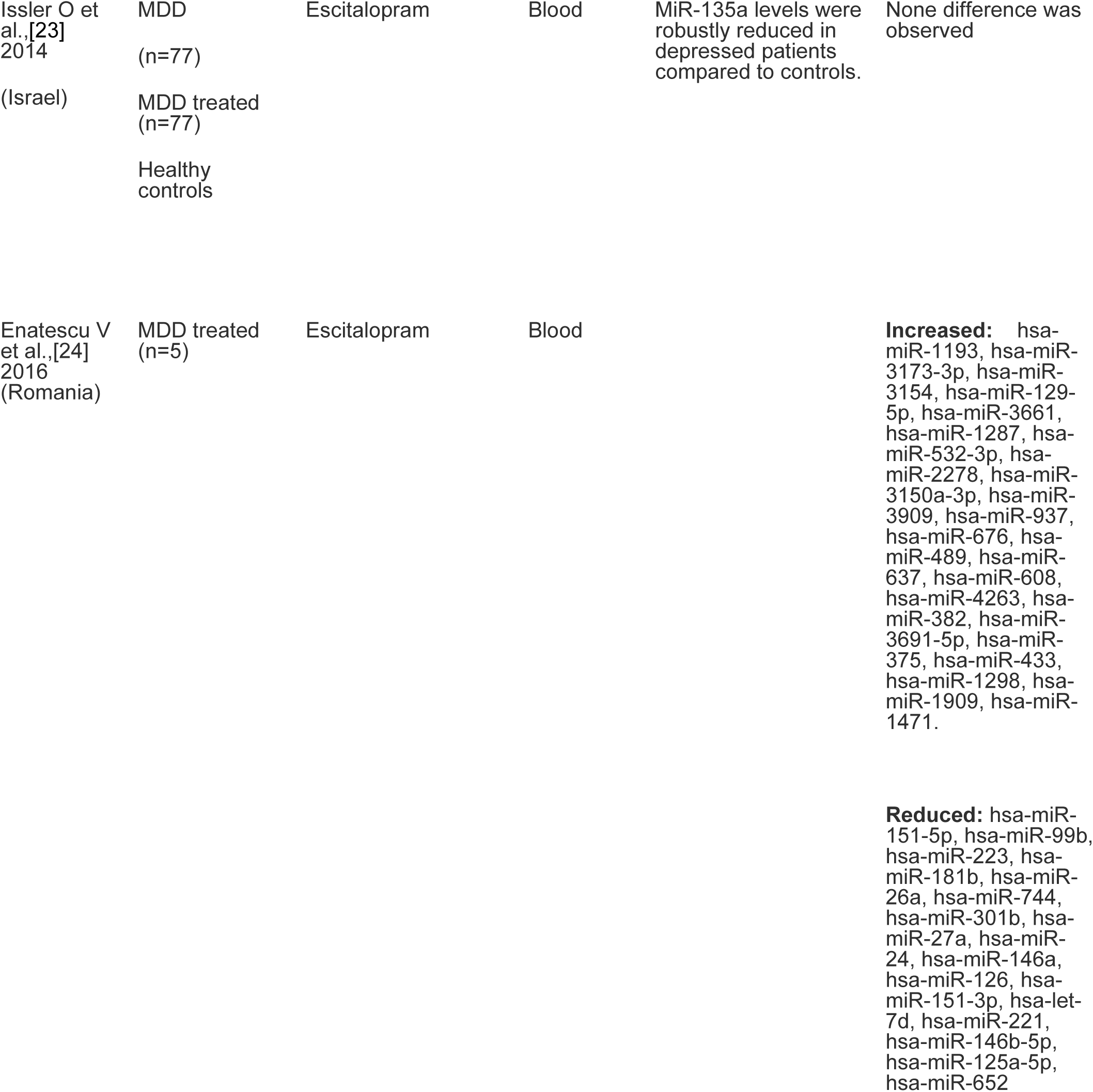
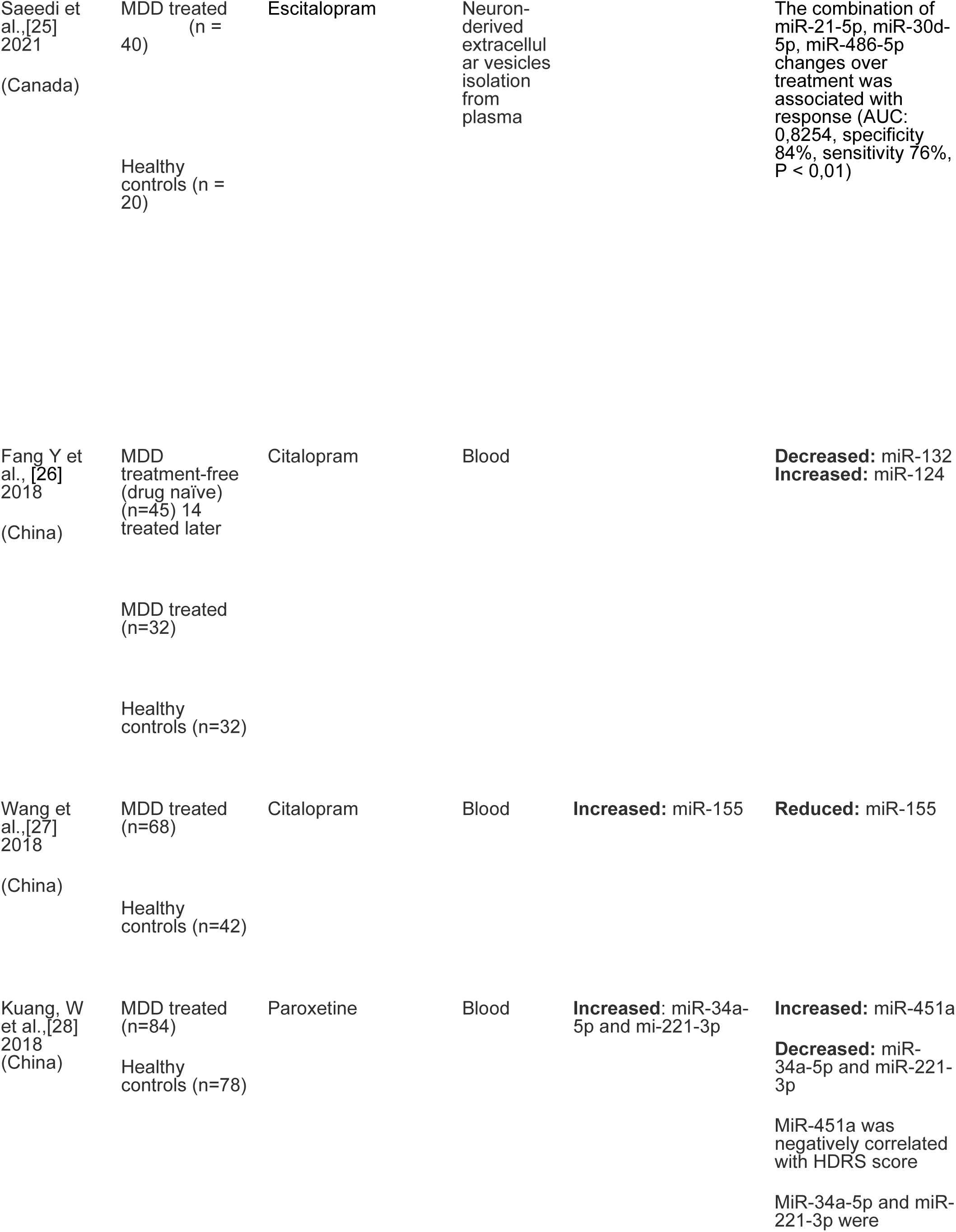
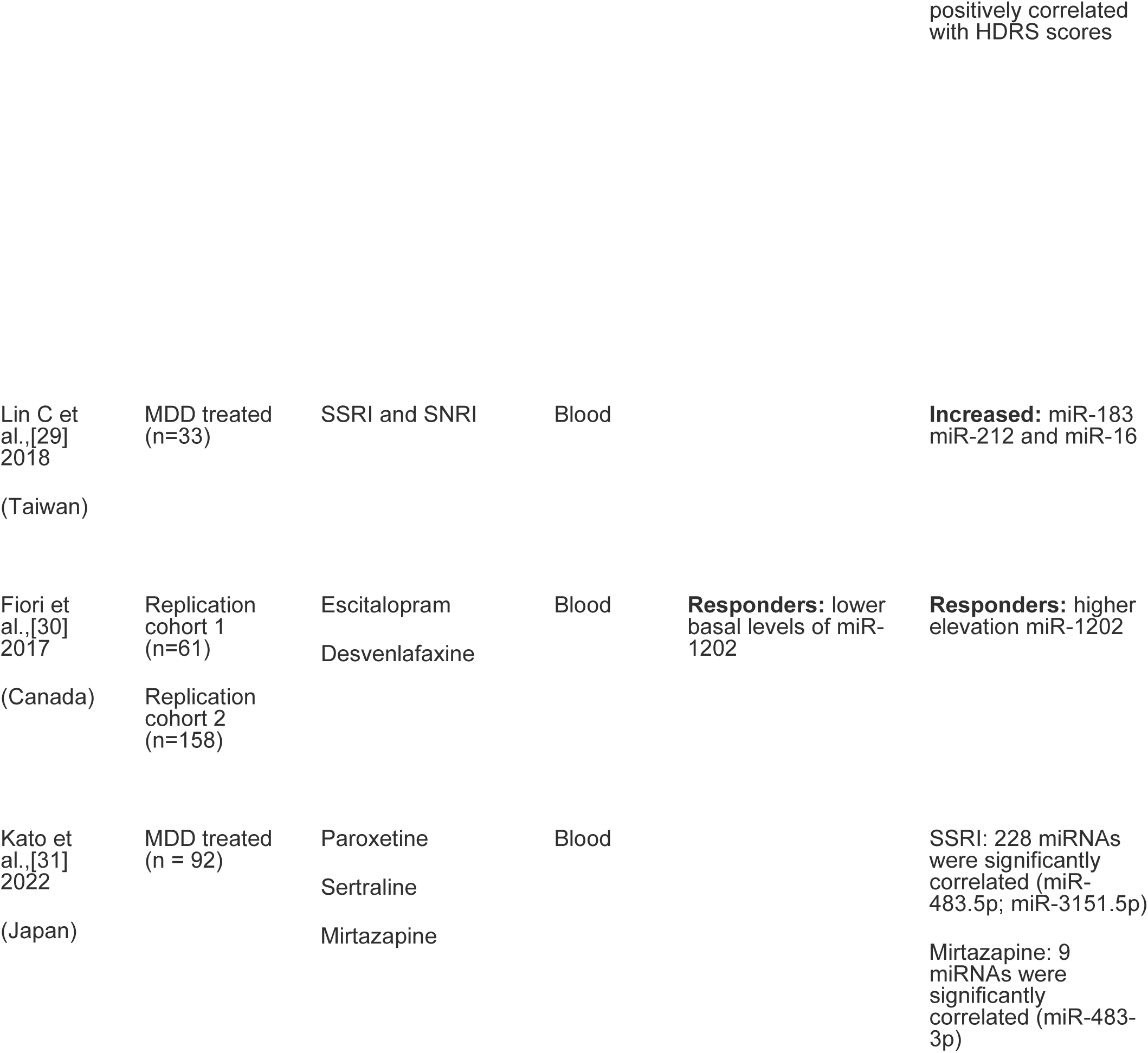
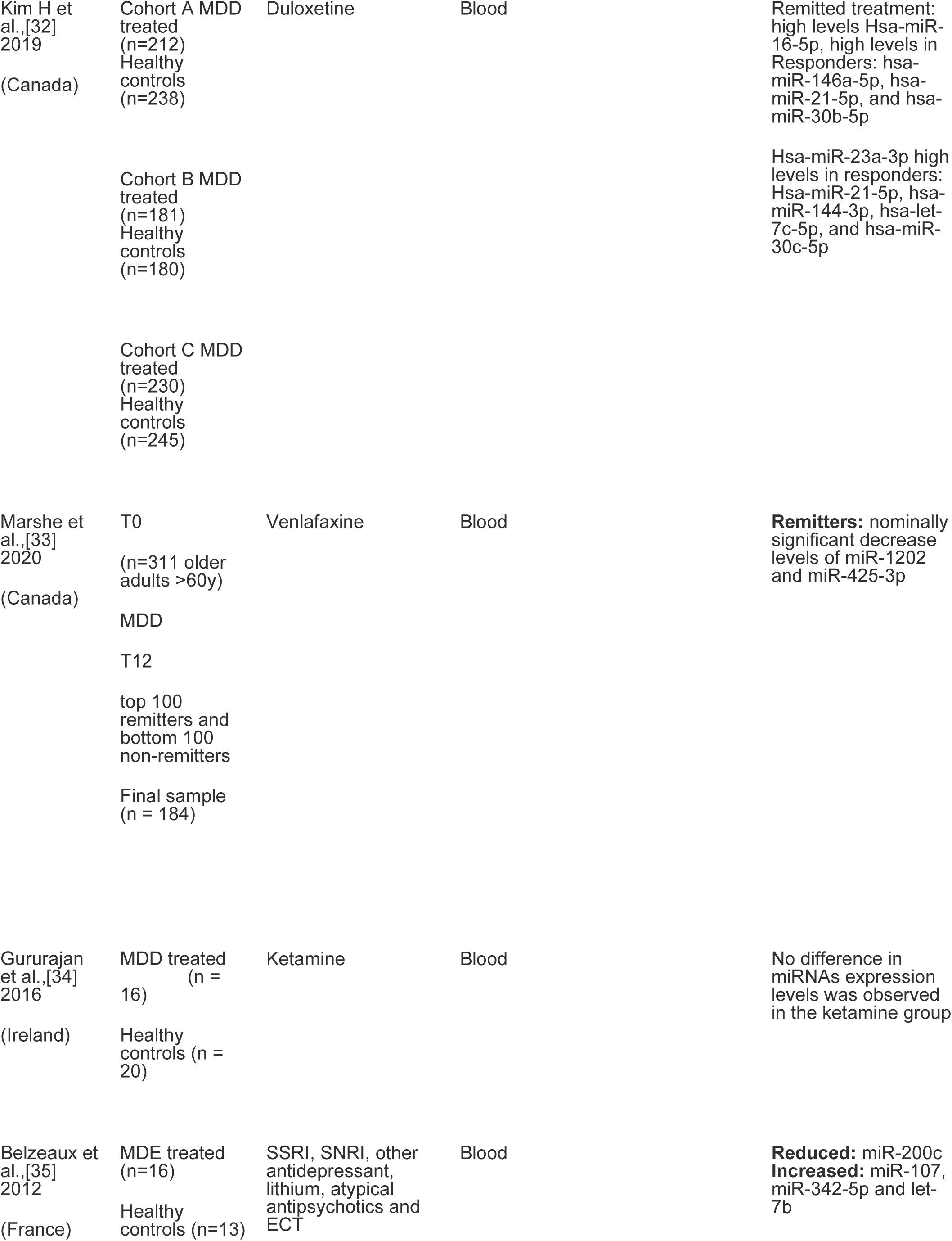
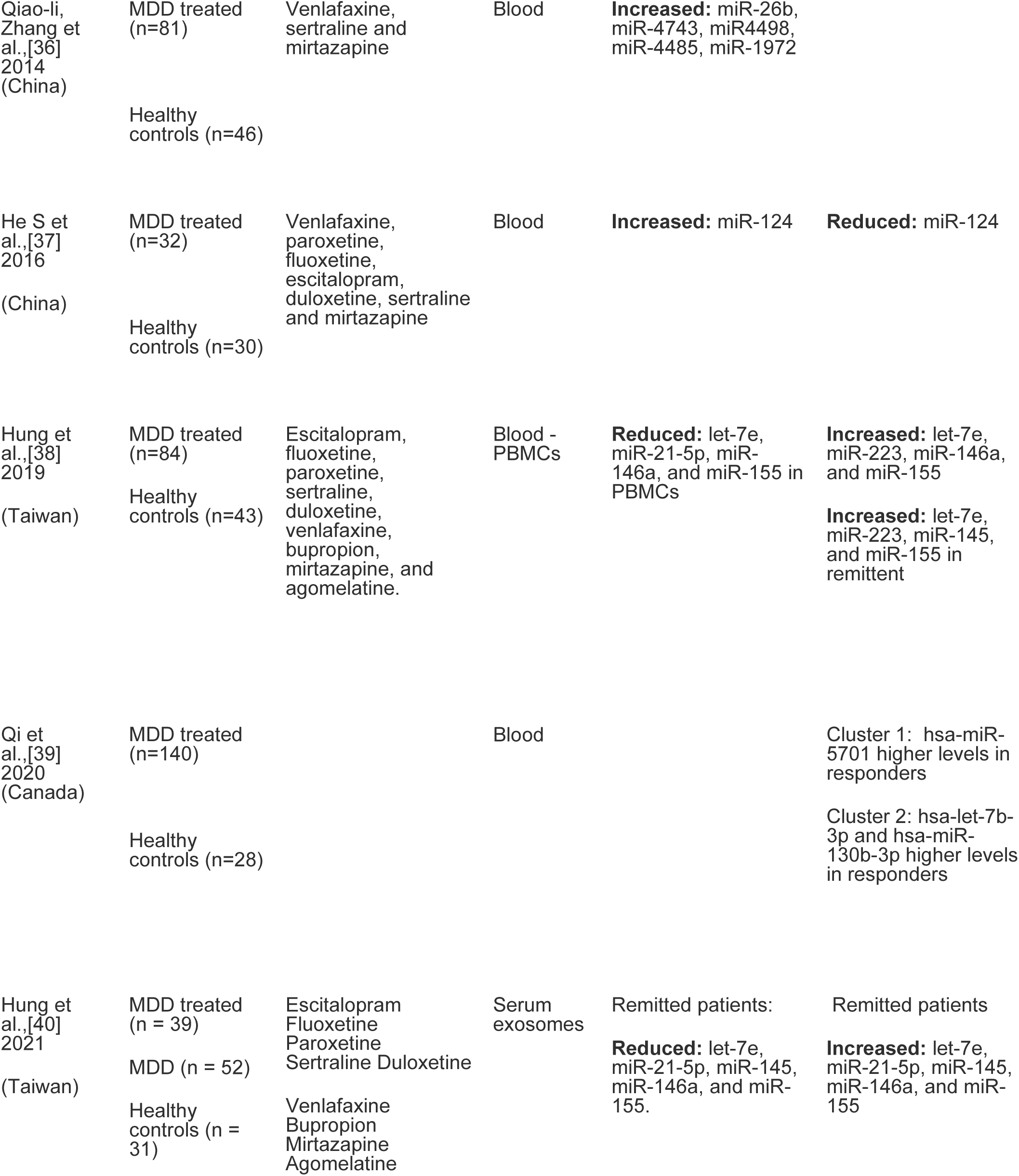
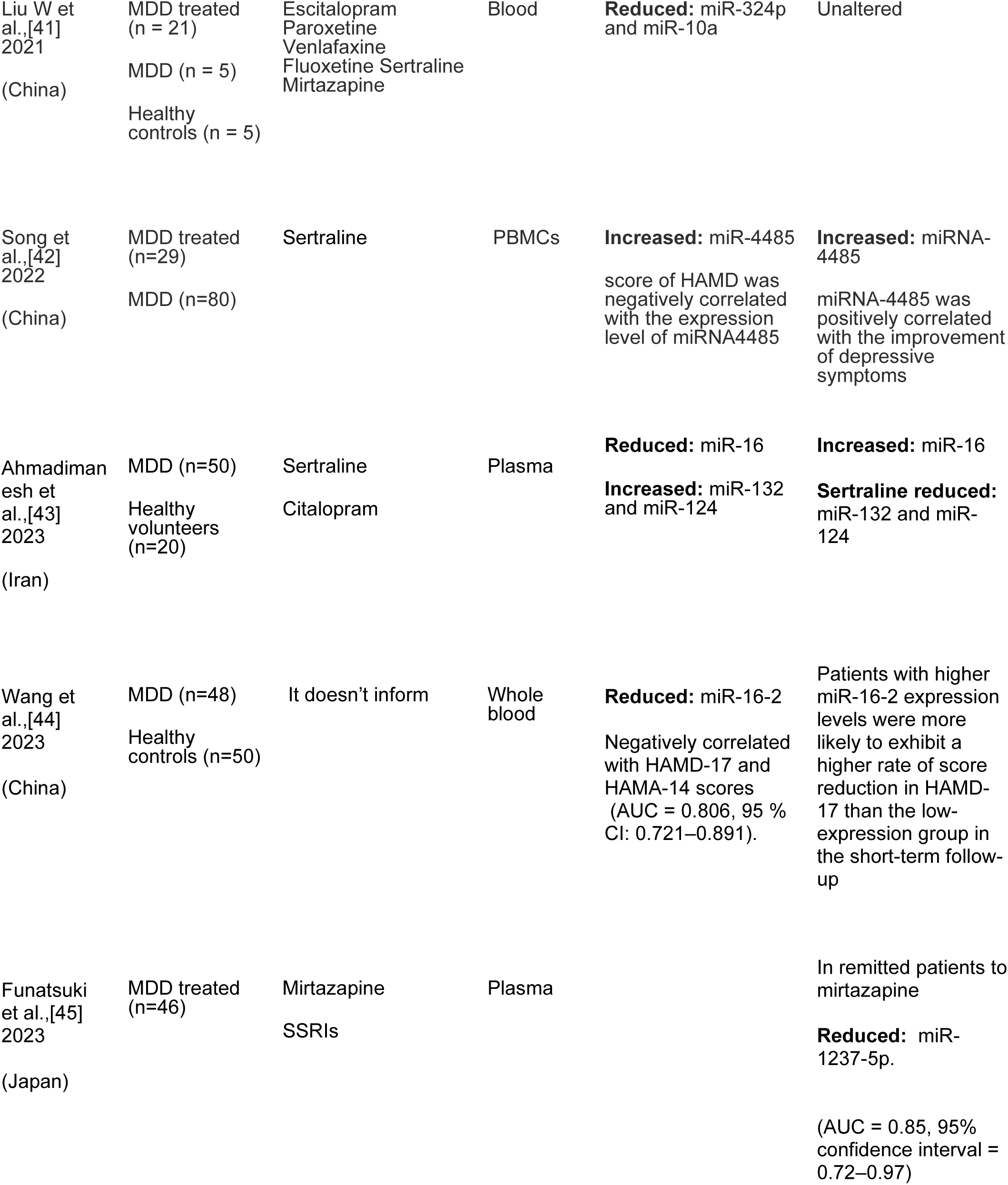
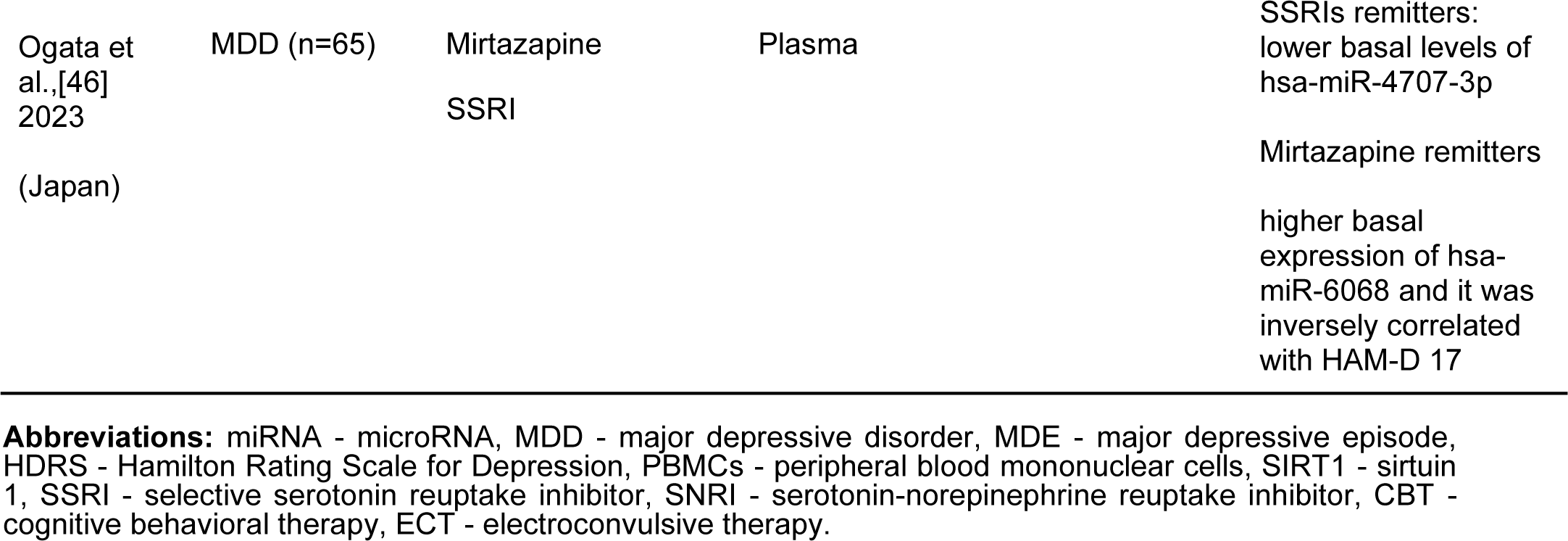
Summary of 28 studies investigating miRNAs expression in humans after MDD treatment.

In 2013, Bocchio-Chiavetto et al. [21] determined that after 12 weeks of treatment with escitalopram, 28 miRNAs became overexpressed, with fold changes ranging from 2.07 to 4.68, whereas only two miRNAs were highly downregulated (miR-34c-5p and miR-770-5p), with fold decreases of 2.03 × 10-2 and 1.69 × 10-2, respectively [21]. Homorogan et al. [22] found that miR-494 was differentially expressed in exosome-depleted plasma after 12 weeks of treatment with escitalopram. Additionally, in a study comparing MDD patients with healthy controls, the MDD group presented lower miR-135a levels in the blood samples taken, and after 12 weeks of escitalopram treatment or cognitive behavioral therapy (CBT), patients undergoing CBT had an increase in miR-135a expression [23]. These effects were not significant in the tests with miR-16 [23]. Furthermore, the brain miR-135a and miR-16 levels of suicide victims were significantly lower than those of the controls, specifically in the dorsal raphe and raphe magnus subnuclei [23]. Enatescu et al. [24] assessed the peripheral expression changes of 222 miRNAs in 5 MDD patients treated with escitalopram for 12 weeks and found that 23 were upregulated and that many of these presented a greater than 2.5 times increases, while 17 of the miRNAs were downregulated [24].

Saeedi et al. [25] found that the combination of changes in miR-21-5p, miR-30d-5p, and miR-486-5p with escitalopram treatment was associated with treatment response (AUC: 0.8254, specificity 84%, sensitivity 76%, P < 0.01). This was the only study that analyzed the specificity and sensitivity of a cluster of microRNAs and highlighted them as good biomarkers of the response to AD treatment with escitalopram. Moreover, when comparing the miR-132 and miR-124 plasma changes among MDD patients that were drug naïve, those that used citalopram, and a control group of individuals without depression, those authors found that miR-132 levels were 2.4 times higher in the drug naïve group when compared to the control, while MDD patients undergoing citalopram had similar miR-132 levels to those of the control group [26]. However, both MDD groups showed higher miR-124 plasma levels than the control group (1.8-fold and 4-fold in the drug naïve and citalopram-treated groups, respectively) [26]. After two months of treatment with citalopram, a decreasing trend in plasma miR-132 and an increase in miR-124 was detected in the 14 drug naïve patients tested [26]. Another study indicated that miR-155 levels were downregulated after treatment with citalopram [27], whereas both cellular and serum levels of miR-155 were found to be significantly upregulated among MDD patients compared to those of healthy controls [27]. Kuang et al. [28] found decreased levels of miR-451, and higher miR-34a-5p and miR-221-3p levels in depressed patients pre- and post-paroxetine treatment compared to the healthy control group. After treatment with paroxetine, miR-451a levels increased, although both miR-34a-5p and miR-221-3p levels decreased significantly when compared to the basal levels [28].

Participants treated solely with SSRIs presented increased levels of miR-16, while patients treated with either SSRIs or serotonin and norepinephrine reuptake inhibitors (SNRI) exhibited increased expressions of miR-183 and miR-212 [29]. Low baseline levels of miR1202 were associated with improved therapeutic responses to SSRIs and SNRIs; MDD patients who responded to AD treatment had lower basal levels of miR-1202, and these levels increased after the drug was administered [30]. Moreover, the prediction analyses of nonresponse found a sensitivity of 91.7%, a specificity of 67.7%, and an AUC=0.812 [30]. Among patients treated with an SSRI (paroxetine or sertraline), a significant correlation was identified two weeks after treatment initiation between 228 miRNAs (especially miR-483-5p and miR-3151-5p) and depression severity among patients treated with mirtazapine, with the strongest association found with miR-483-3p [31].

Three studies assessed miRNA levels in patients treated with duloxetine, venlafaxine, or ketamine. For patients treated with duloxetine, baseline levels of hsa-miR-23a-3p were associated with a remission of depressive symptoms [32]. Belzeaux et al. [11] reported elevated baseline levels of miR-135a-5p and miR-16 and an improved treatment response in patients administered citalopram. Patients whose depressive symptoms abated had nominally significant decreases in miR-1202 and miR-425-3p levels [33]. Among the patients who received ketamine, there were no differences in miRNA expression when compared to the control group [34]. Other studies evaluated patients who received different ADs. Belzeaux et al. [35] found that after the treatment of patients experiencing a major depressive episode, miR-107, miR-342-5p, and let-7b levels were upregulated, while miR-200c expression was downregulated [35]. Depressed patients showed significantly higher expressions of miR-26b, miR-4743, miR-4498, miR-4485, and miR-1972 before treatment [36].

Various studies have investigated the basal characteristics of patients in the search for treatment response predictors and the untreated MDD miRNA profile. Peripheral blood mononuclear cells (PBMCs) showed elevated levels of miR124 in patients with depression, and a reduction in these levels was associated with improved treatment responses [37]. After adjustments for age, sex, smoking status, and body mass index (BMI), the levels of let-7e, miR-21-5p, miR-146a, and miR-155 were significantly lower in PBMCs isolated from baseline MDD patients [38], while let-7e and miR-146a were negatively correlated with the HAMD-17 score after treatment, and miR-155 was positively correlated with this value [38]. Qi et al. [39] used machine-learning analyses of miRNA to reveal high hsa-miR-5701 levels among responders in one cluster and high hsa-let-7b-3p and hsa-miR-130b-3p levels in a second cluster [39]. Reduced levels of miR-146b-5p, miR-24-3p, and miR-425 were associated with better responses to treatment in both human and animal models [12]. Hung et al. [40] showed that patients who achieved remission after treatment with the different antidepressants had lower basal levels of let-7e, miR-21-5p, miR-145, miR-146a, and exosomal miR-155 compared to those of healthy controls, which was followed by a notable increase in these levels post-treatment. Although most studies showed changes in miRNA levels after treatment, Liu et al. [41] found reduced baseline levels of miR-324b and mir-10a in depressed patients when compared to healthy controls that were unchanged after 8 weeks of treatment with classic antidepressants. The sertraline treatment reduced the high miR-4485 levels shown in MDD patients that were negatively correlated with HAMD scores [42].

Two recently published studies have identified decreased levels of miR-16 in patients diagnosed with MDD, with one study demonstrating an increase in these levels following treatment with sertraline or escitalopram (Ahmadimanesh et al., 2023 [43]; Wang et al., 2023 [44]). Sertraline, but not escitalopram, was found to significantly reduce the levels of miR-132 and miR-124, both of which were elevated in MDD patients (Ahmadimanesh et al., 2023 [43]). Furthermore, according to Wang et al., 2023 [44], it was observed that miR-16-2 exhibited a negative correlation with both HAMD-17 and HAMA-14 scores (AUC = 0.806, 95% CI: 0.721–0.891) (Wang et al., 2023[44]). Patients displaying higher expression levels of miR-16-2 were more inclined to experience a greater reduction in HAMD-17 scores during short-term follow-up compared to the low-expression group (Wang et al., 2023[44]).

The remaining two studies discovered decreased levels of miR-1237-5p in patients who experienced symptom remission with mirtazapine (AUC = 0.85, 95% confidence interval = 0.72–0.97), along with elevated basal expression of hsa-miR-6068 (Funatsuki et al., 2023 [45]; Ogata et al., 2023[46]). Moreover, one of these studies noted a decrease in basal levels of hsa-miR-4707-3p in remitters to SSRIs (Ogata et al., 2023[46]).

### 3.2. Animal models

We found fourteen studies comparing the miRNA expressions of rodents undergoing AD treatment (Table *4*). Three SSRI studies reported diverse miRNA profiles and variable treatment response patterns among the animal samples. Patricio et al. [47] investigated changes in miR-409-5p and miR-411-5p levels in brain regions linked to depression in rats undergoing chronic mild stress (CMS). The rats received treatment with either fluoxetine or a saline solution [47]. The stress-exposed animals presented significantly increased levels of miR-409-5p in the nucleus accumbens that were not reversed with fluoxetine treatment [47]. In the dorsal dentate gyrus, chronic fluoxetine use resulted in increased levels of miR-411-5p; therefore, miR-411-5p levels were significantly decreased in the blood of fluoxetine-treated rats [47]. The genetic deletion of miR-34s impairs the effect of chronic fluoxetine on the behavioral responses of rats to an acute threat [48]. Chronic fluoxetine treatment increased the levels of miR-34a in WT mice, but this was not observed with acute administration of this compound [48].

**Table 4.**
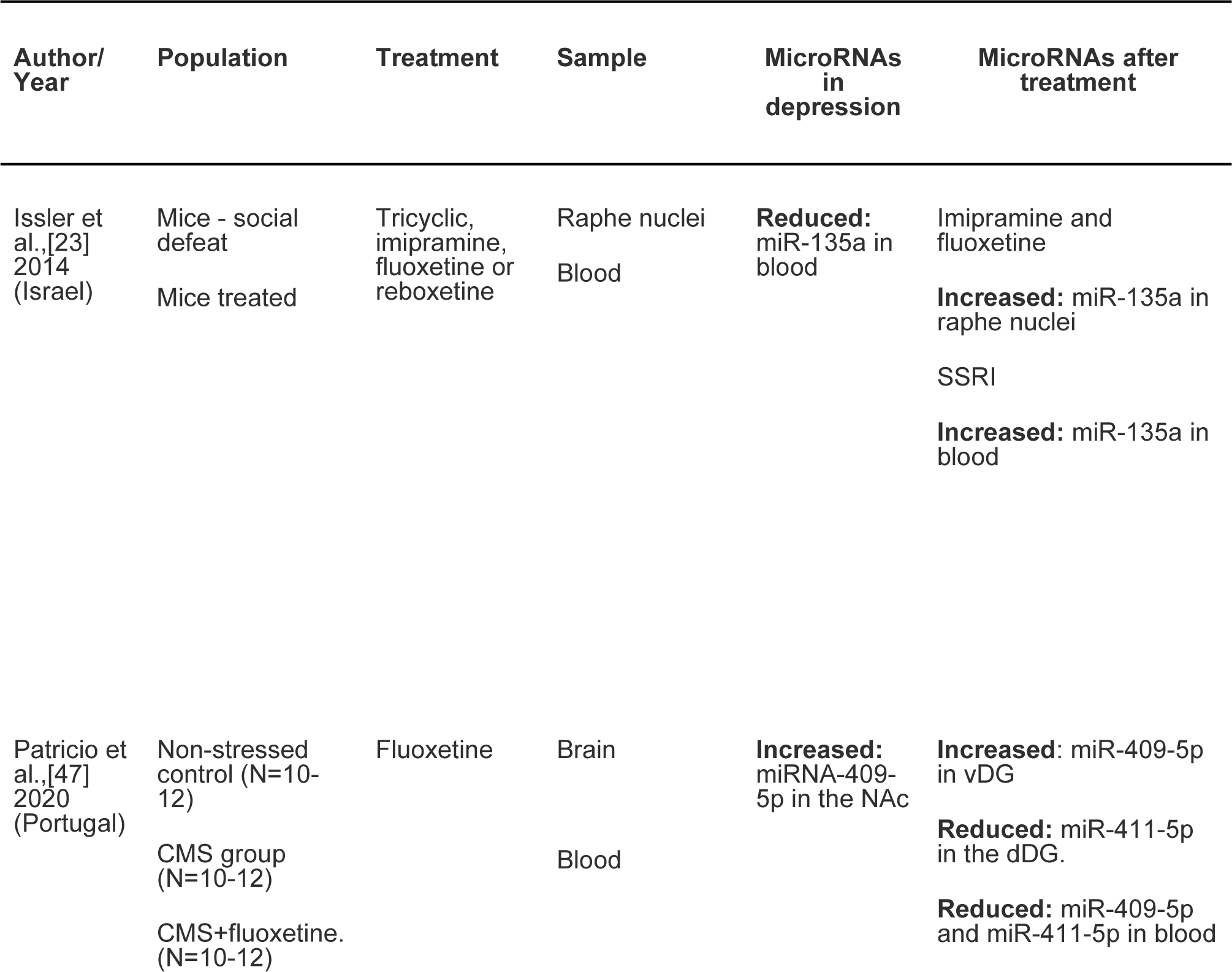
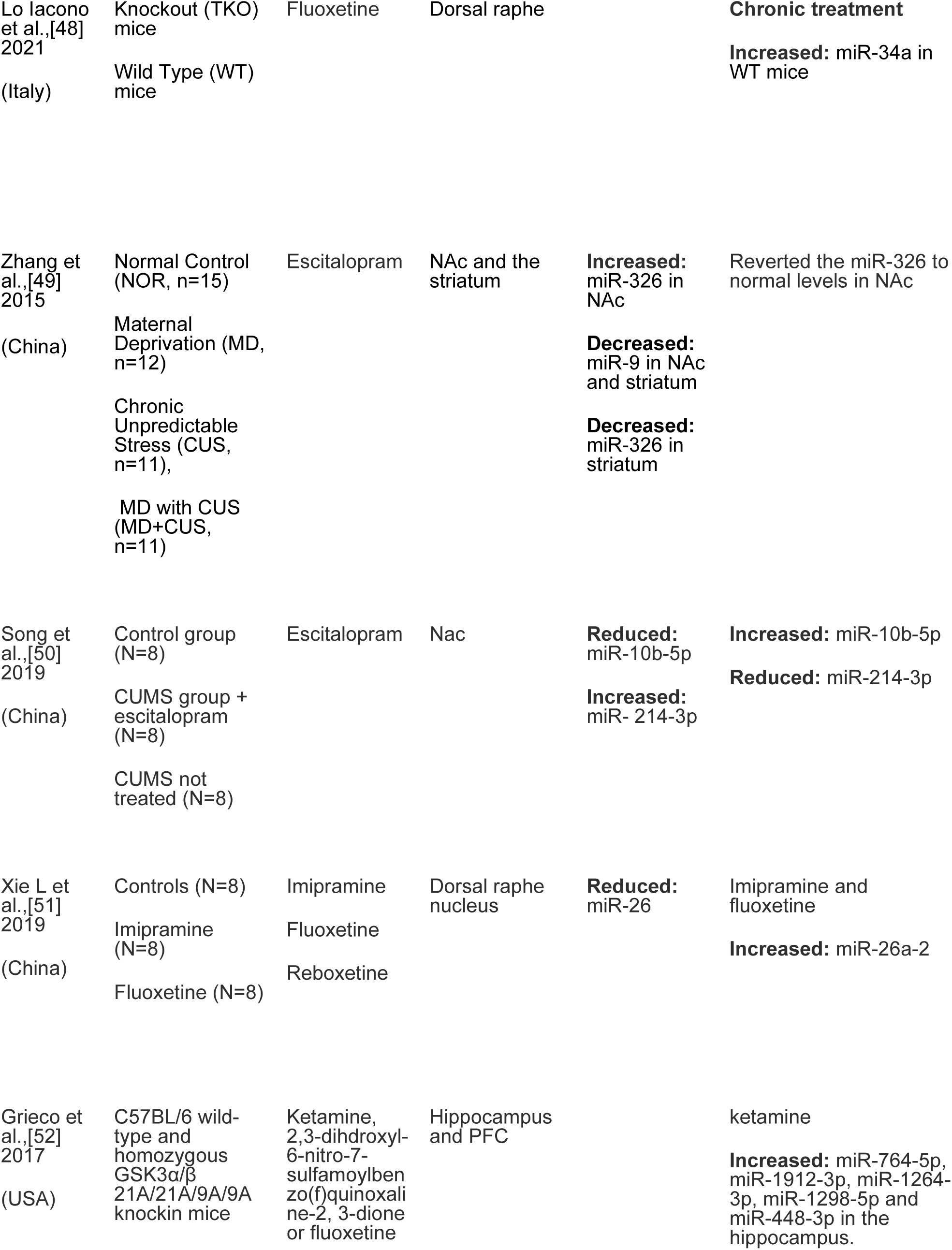
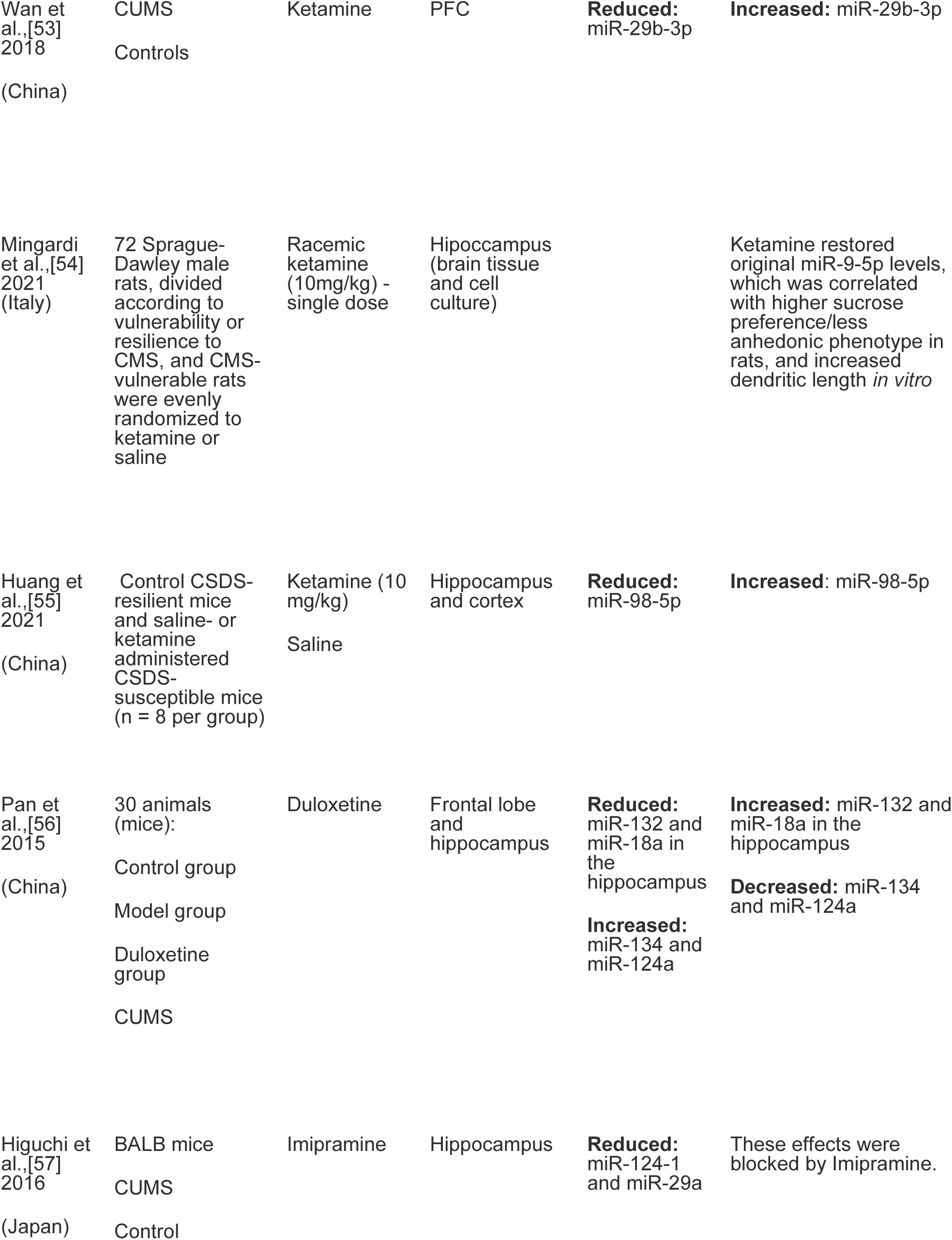
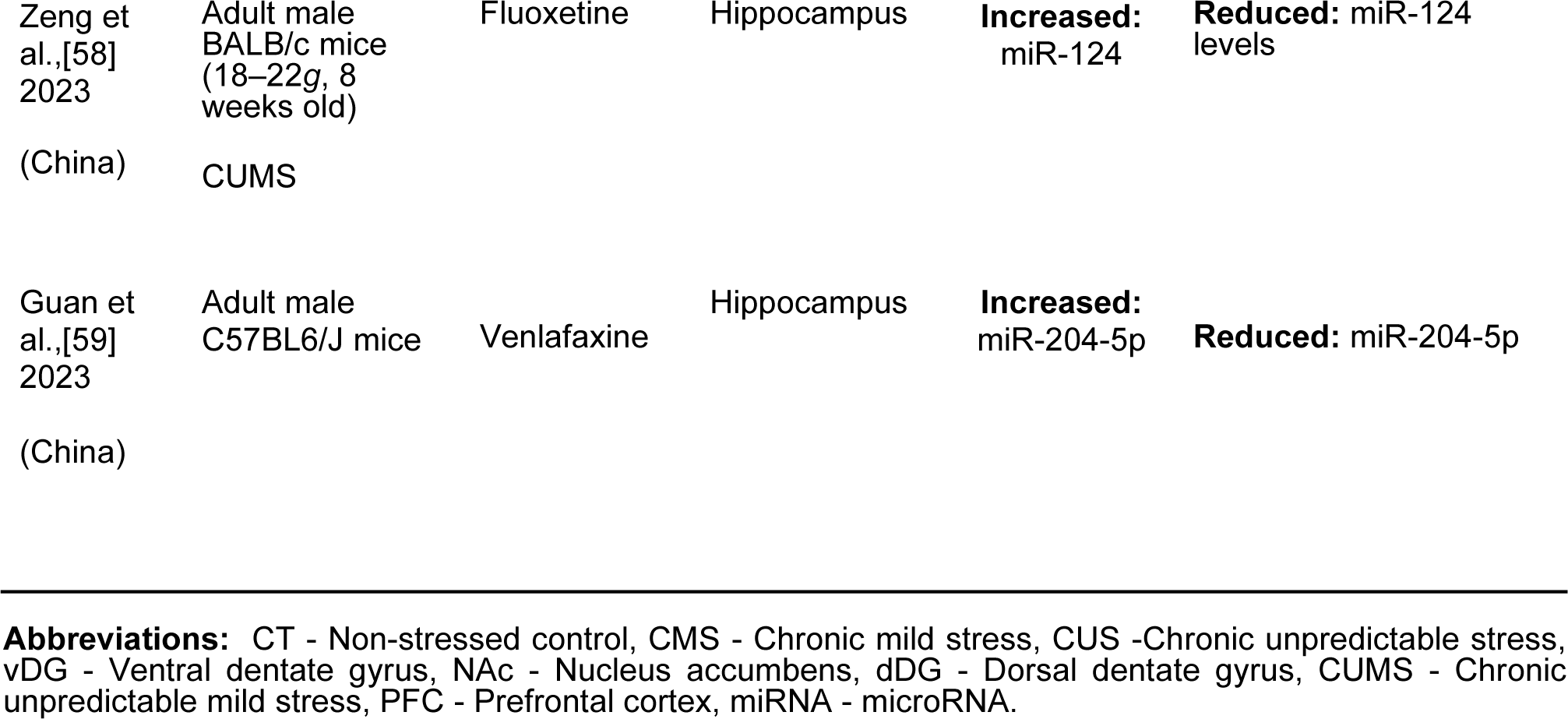
Summary of 14 studies investigating miRNAs expression in animal models after AD use.

Another study investigated the changes in the expression of miR-326 and miR-9 [49]. Adult rats exposed to chronic unpredictable stress (CUS) and maternal deprivation had lower levels of miR-9 in the nucleus accumbens and striatum, and these levels did not rise after escitalopram treatment [49]. In the nucleus accumbens, the altered expression of miR-9 was negatively correlated with an overexpression of the dopamine receptor D2 (DRD2) protein, which appears to be involved in chronic stress and early-life adversity [49]. The expression of miR-326 was increased in the nucleus accumbens and decreased in the striatum of rats exposed to CUS [49]. In addition, animals with maternal deprivation showed increased levels of miR-326 in the striatum [49]. In contrast to the miR-9 findings, miR-326 levels returned to normal after escitalopram treatment [49]. In another study, escitalopram treatment reversed miRNA expression alterations in rats after chronic unpredictable mild stress (CUMS) [50]. Although those authors found 18 miRNA alterations, they suggested that miR-10b-5p and miR-214-3p had important contributions to the miRNA network of the nucleus accumbens since the expression of these miRNAs differed significantly between the stressed group and the control, and between the treated group and the control [50].

The miRNAs related to serotoninergic networks were influenced by AD treatments in some studies. A potential regulator of serotoninergic activity, miR-135, was increased after AD treatment [23]. High levels of miR-135a appear to diminish depression-like behaviors after stress exposure, whereas its absence reduces the response to ADs [23]. A study using transgenic mice suggested that miR-26a-2 levels are involved in the serotoninergic network, given that ADs upregulate the expression of this miRNA in the dorsal raphe nucleus [51].

The influence of ketamine on miRNA expression was evaluated, and one study reported variable changes in the expression of miRNA serotonergic (5HT)-2C-receptor (5HTR2C) clusters in mouse hippocampi [52]. Following 24 hours of ketamine infusion, miR-764-5p, miR-1912-3p, miR-1264-3p, miR-1298-5p, and miR-448-3p were upregulated [52], with miR-448-3p showing the greatest increase and poorest AD response when it was inhibited. [52]. These changes were not replicated in the prefrontal cortex [52]. The same miRNA changes were not observed in mice receiving fluoxetine, which suggests that these are part of a ketamine response and not just a typical AD response [52]. In addition, miR-29b-3b appears to be involved in the AD effect of ketamine. A study found that this miRNA level was reduced in the prefrontal cortex of rats after CUMS; however, after ketamine treatment, the miR-29b-3b levels were reestablished [53].

Another study found that the administration of a single dose of ketamine in CMS rats restored the original miR-9-5p levels, and this increase was correlated with higher sucrose preference/less anhedonic phenotype [54]. In addition, Huang et al. [55] explored the antidepressant effect of ketamine in an animal model of depression and observed a reduction in miR-98-5p levels in the prefrontal cortex and hippocampus after treatment.

In a study with mice undergoing CUMS, duloxetine was found to enhance the expression of miR-18a in the frontal lobe, as well as upregulate miR-132 and miR-18a and downregulate miR-134 and miR-124a in the hippocampus [56].

Higuchi et al. [57] found lower levels of miR-124 in the hippocampus of mice subjected to chronic stress. The upregulation of miR-124 in the hippocampus increased resilience in mice, while its downregulation enhanced depression-like behaviors [57]. In other hand, Zeng et al., 2023 [58] found increased level of miR-124 in the hippocampus of mice with symptoms like depressive and the fluoxetine administration reduced these levels. Guan et al., 2023 [59] investigou os níveis do miR-miR-204-5p em ratos com fenótipo tipo depressivo e possíveis alterações causadas pelo tratamento com a velanfaxina. The group found higher levels of miR-204-5p in depression and the treatment.reduced this levels.

### 3.3. Cell cultures

We included four cell culture studies targeting different miRNAs and a variety of sample cultures (Table *5*). Three of these used paroxetine and one used duloxetine.

**Table 5.**
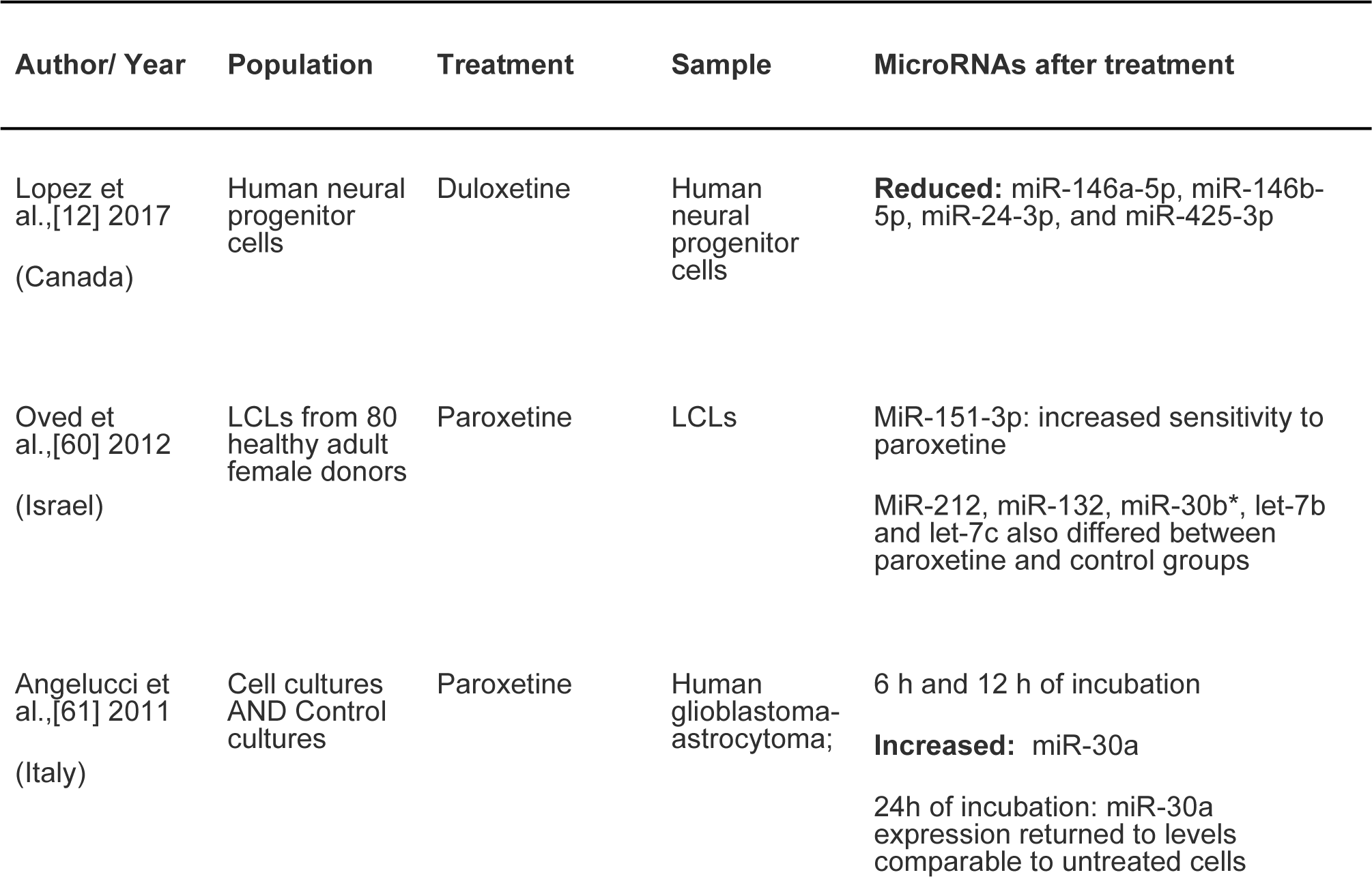
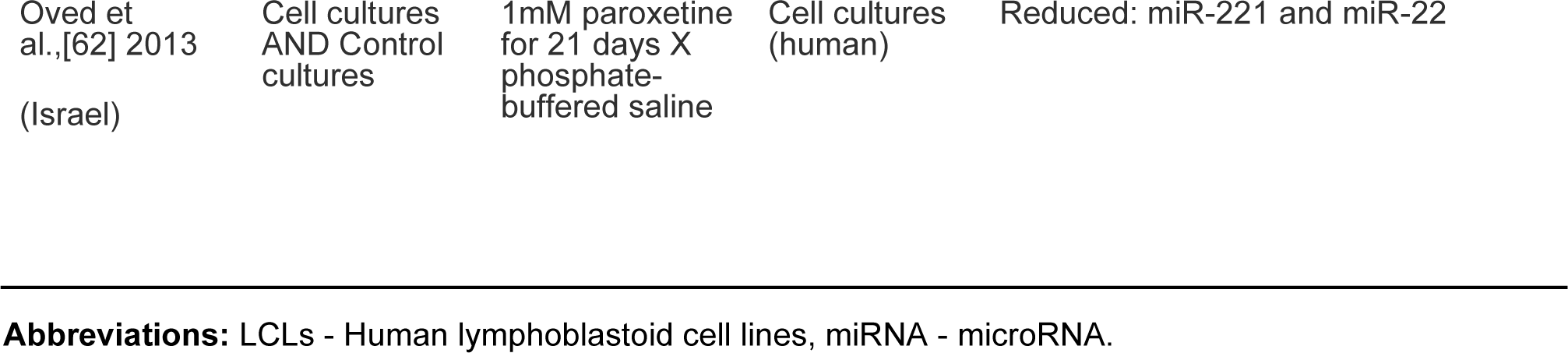
Summary of 4 studies investigating miRNAs expression in human cell cultures after ADs use.

The exposure of human lymphoblastoid cell lines to paroxetine for three days increased the expression of miR-212, miR-132, miR-30b, let-7b, and let-7c [60], and its incubation for 21 days decreased the expression of miR-221 and miR-22.55. The first study identified a correlation that mainly concerned the miR-151-3p basal levels, which were higher in cells that were sensitive to the paroxetine treatment [60]. Another study by the same group primarily showed a downregulation of miR-221 and miR-222 after treatment [61]. The third study using paroxetine analyzed glioblastoma-astrocytoma cell line U87 and found a significant upregulation of miR-30a levels beginning 6 hours after treatment, which lasted for 12 hours after treatment [62]. The final study used duloxetine as the treatment, and this was previously discussed in the human studies section of this article. Those authors confirmed their previous findings on human and rodent studies in an in vitro analysis using human neural progenitor cells, with reduced levels of miR-146a-5p, miR-146b-5p, miR-24-3p, and miR-425-3p in the cell culture [12].

## 4. Discussion

We found several studies that evaluated miRNA expression in humans, animals, and cell cultures, both before and after receiving ADs. Different classes of AD were used, as well as a broad profile of expressed miRNAs.

The studies revealed altered levels of specific miRNAs in patients with MDD before and after receiving treatment, which indicates that miRNAs could be involved in the pathophysiology of MDD and response to ADs, and be potential biomarkers for MDD diagnosis and therapeutic response. The studies evaluated were heterogeneous, with no pattern of change observed in the miRNAs; therefore, more robust studies with larger samples and consistent methodologies are required.

Most of the microRNAs investigated in the studies are suggested to be involved in the mechanisms of neuroplasticity, such as in the expression of brain-derived neurotrophic factor (BDNF), which is an important neurotrophin that is possibly involved in MDD pathophysiology and treatment response to ADs such as miR-1202, miR-124, miR-16, and miR-135 [57,63]. Wibrand et al. [63] reported that the overexpression of miR-34a was linked to a reduction in the BDNF.

Another microRNA that may be important for the pathophysiology of depression and AD response is miR-124 [57], which could have a central role in neurogenesis and neuronal function, as well as in regulating proliferation, growth, and apoptosis of the central nervous system (CNS) [26,37,64]. Mir-124 has also been shown to negatively affect BDNF expression [37,65]. Lin Sheng Shi et al. [66] provided evidence supporting the role of miR-124 in depression-induced mice, which had increased depression-like behaviors and higher miR-124 expression levels in the hippocampus than the control mouse. In addition, those authors found that the genetic knockdown of hippocampal miR-124 exerted significant antidepressant-like effects that were mediated by the promotion of BDNF biosynthesis. These results corroborate the findings presented in other articles regarding the importance of miR-124 in the physiopathology of depression and antidepressant response.

Mir-132 has been linked to important and critical CNS functions, including neurogenesis and synaptic plasticity in different brain regions [26,67]. MiR-212, miR-30b, let-7b, and let-7c have been identified as relevant to the neuroplasticity process, while miR-16, miR-183, miR-212, and miR-221-3p are related to BDNF expression [28,29,60,61].

Lopez et al. [12] proposed a mechanism of action for miR-146a/b-5, miR-425-3p, and miR-24-3p as regulators of the MAPK and Wnt pathways based on the results of studies conducted on humans and cell cultures. These miRNAs regulate a range of genes involved in the signaling process of the two pathways, in addition to interfering with the BDNF pathway. The genes regulated by these miRNAs have been linked to MDD and AD response. Other relevant microRNAs are miR-34c-5p and miR-34a-5p, which are suggested to be related to stress response and neuroplasticity [21,28], while the genetic deletion of the miR-34s can hinder the antidepressant effect [48].

MiR-135a is involved in the metabolism and level of 5-HT, whereby it is an important regulator for maintaining serotonergic tone to centrally favor the AD response [23]. MiR-135a-3p is involved in the modulation of the glutamatergic system and stress response, although its association with these processes is unclear [11,33].

Other microRNAs, such as let-7e, miR-21-5p, miR-146a, and miR-155, have been related to inflammatory processes that are associated with important pathways for the pathophysiology of MDD. In addition, two studies focusing on brain tissue miRNAs from MDD patients have identified the role of miR-1202 in regulating the glutamate receptor, which is a new pathway that is proposed to be involved in MDD. MiR-1202 is a primate-specific miRNA that is associated with the pathophysiology of MDD and the responsiveness to selective serotonin reuptake inhibitors (SSRIs) [68]. The potential of miRNAs as biomarkers for response to treatment has been indicated [69]. MiR-1202 regulates metabotropic glutamate receptor 4 (GRM4), which is a proposed target glutamatergic receptor for the development of ADs. AD treatment was shown to increase miR-1202 levels and decrease GRM4 levels. Initial miR-1202 expression was lower in responders to ADs, while non-responders demonstrated no change in miR-1202 expression compared to healthy individuals [68].

The findings of this systematic review reveal that miRNAs have been linked to MDD and AD treatment. This knowledge can contribute to the understanding of depression etiology and antidepressant action. Based on these findings, miRNA expression can be used to direct the treatment of MDD [70]. Thus, this is the first review to explore the changes in miRNAs after AD exposure in three different populations (cell cultures, animal models, and human studies).

### 4.1. Limitations

The limitations of our study include the diversity of the populations studied (humans, animal models, and cell cultures), wide range of ADs used, and large number of miRNAs identified. Although a wide variety of miRNAs exist, there is little replication of studies that investigate the same molecules from different perspectives. Most studies are limited to evaluating the change of miRNAs in AD treatments, without assessing or describing the related action mechanisms. These aspects complicate meta-analyses; therefore, a discussion and understanding of the basic aspects of the pathophysiology of depression are limited. Although the literature has established a relationship between circulating miRNAs and MDD, it is important to understand the targets of these miRNAs using in vitro, in vivo, and in silico models.

The studies included in this review assessed specific miRNAs that were associated with MDD and AD response. They provide evidence that specific miRNAs can act as biomarkers; however, the clinical utility of miRNAs as biomarkers of MDD and responses to ADs needs to be further investigated.

## 5. Conclusions

The results of our systematic review demonstrated that certain microRNAs are altered in patients with MDD and can be modified with AD treatment, corroborating their potential as diagnostic and response biomarkers. Understanding the pathophysiology of MDD can improve the therapeutic options for this condition. Considering the heterogeneity of miRNAs across the studies, further investigation is required to evaluate the expressions of miRNAs in MDD and their changes with treatment to confirm these findings and provide a comprehensive explanation of the involved pathways. Existing evidence supports the hypothesis that miRNAs could have a place in the future of personalized psychiatry.

## Data Availability

All data produced in the present work are contained in the manuscript

---

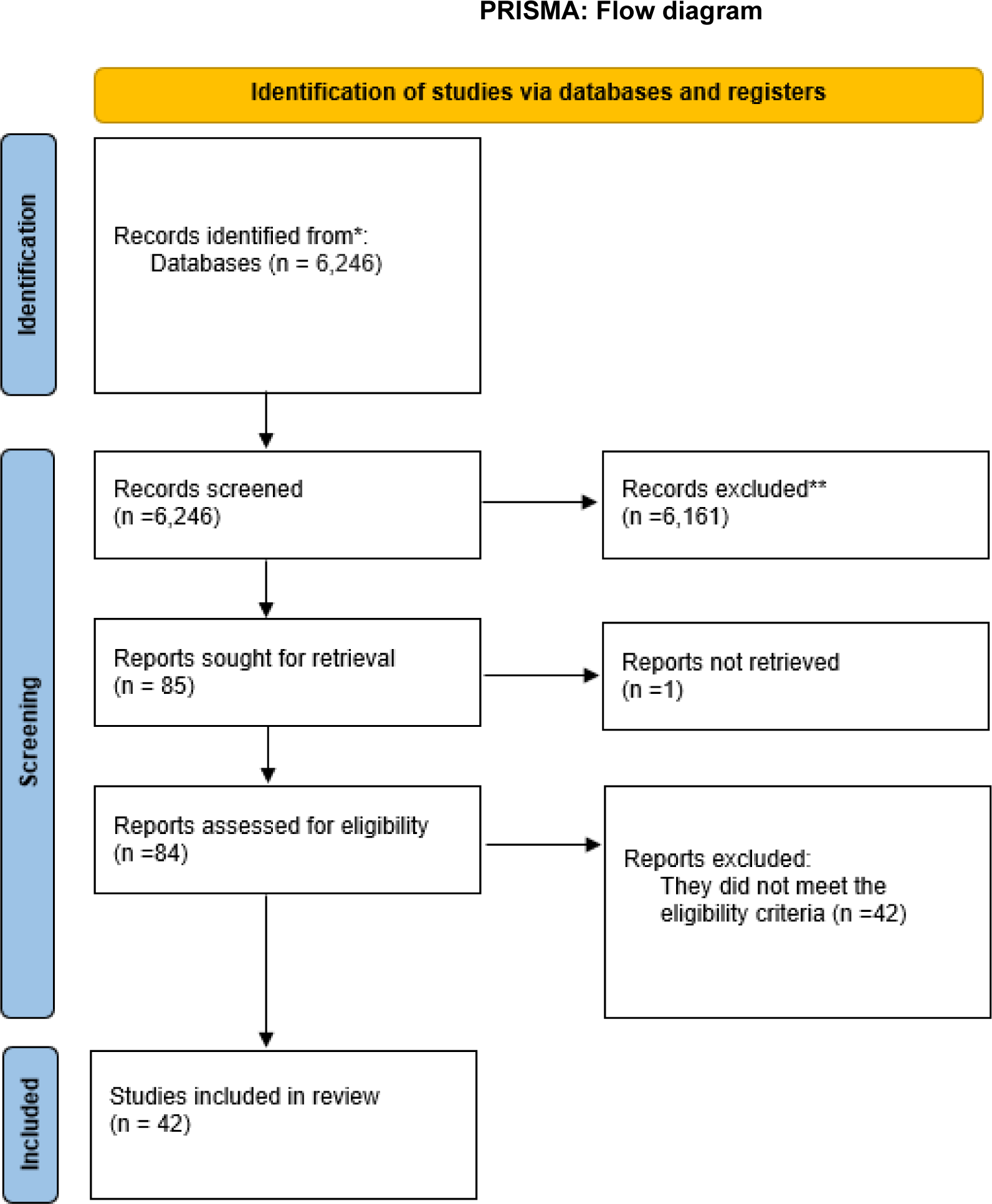

**Table 4.**
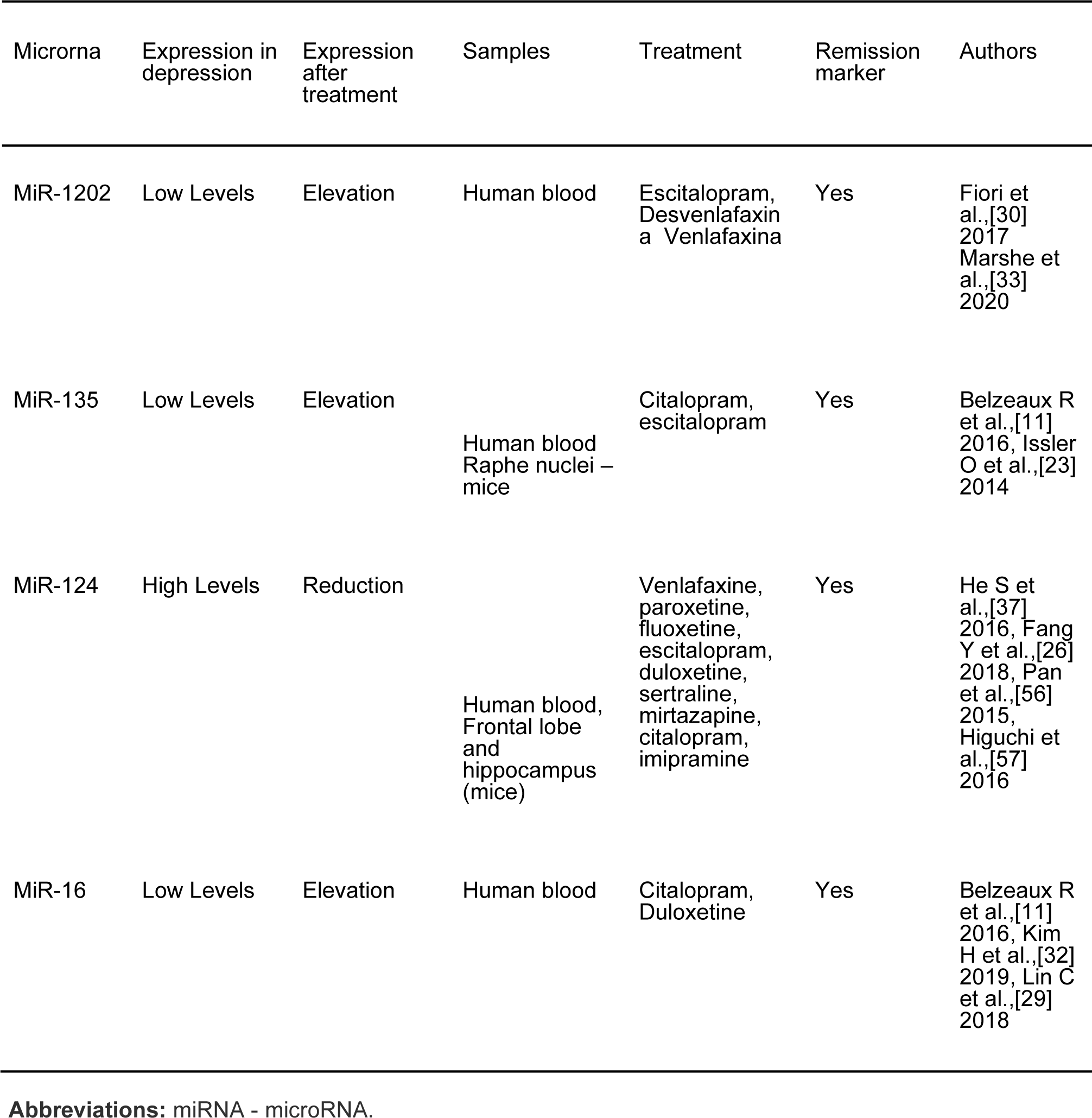
Most consistently expressed miRNA.

## References

1. Belmaker RH, Agam G: Major Depressive Disorder. New England Journal of Medicine. 2008, 358:55–68. 10.1056/NEJMra073096

2. Bromet E, Andrade LH, Hwang I, et al.: Cross-national epidemiology of DSM-IV major depressive episode [Internet]. 2011.

3. Warden D, Rush AJ, Trivedi MH, Fava M, Wisniewski SR: The STAR*D Project Results: A Comprehensive Review of Findings. 2007, Available from: http://www.star-d.org:10.1007/s11920-007-0061-3

4. Tansey KE, Guipponi M, Hu X, et al.: Contribution of Common Genetic Variants to Antidepressant Response. Biol Psychiatry. 2013, 73:679–82. 10.1016/j.biopsych.2012.10.030

5. Leistedt SJ, Linkowski P: Brain, networks, depression, and more. European Neuropsychopharmacology. 2013, 23:55–62. 10.1016/j.euroneuro.2012.10.011

6. Marsden WN: Synaptic plasticity in depression: Molecular, cellular and functional correlates. Prog Neuropsychopharmacol Biol Psychiatry. 2013, 43:168–84. 10.1016/j.pnpbp.2012.12.012

7. Ota KT, Duman RS: Environmental and pharmacological modulations of cellular plasticity: Role in the pathophysiology and treatment of depression. Neurobiol Dis. 2013, 57:28–37. 10.1016/j.nbd.2012.05.022

8. Kanherkar RR, Bhatia-Dey N, Csoka AB: Epigenetics across the human lifespan. Front Cell Dev Biol. 2014, 2:10.3389/fcell.2014.00049

9. Uchida S., Yamagata H., Seki T., et al.: Epigenetic mechanisms of major depression: Targeting neuronal plasticity. Psychiatry. 2018, 72:212–227. 10.1111/pcn.12621

10. Lopez, J. P., Kos, A., & Turecki, G. (2018: Major depression and its treatment: microRNAs as peripheral biomarkers of diagnosis and treatment response. Current. 31:7–16. 10.1097/YCO.0000000000000379

11. Belzeaux R., Lin C., Ding Y., et al.: Predisposition to treatment response in major depressive episode: a peripheral blood gene coexpression network analysis. Journal of psychiatric research. 2016, 81:119–126. 10.1016/j.jpsychires.2016.07.009

12. Lopez JP, Fiori LM, Cruceanu C, et al.: ARTICLE MicroRNAs 146a/b-5 and 425-3p and 24-3p are markers of antidepressant response and regulate MAPK/Wnt-system genes. Nat Commun [Internet. 20178, 10.1038/ncomms15497

13. Dwivedi Y: Evidence demonstrating role of microRNAs in the etiopathology of major depression. J Chem Neuroanat. 2011, 42:142-56. 10.1016/j.jchemneu.2011.04.002

14. Im HI, Kenny PJ: MicroRNAs in neuronal function and dysfunction. Trends Neurosci. 2012, 35:325–34. 10.1016/j.tins.2012.01.004

15. Garza-Manero S, Pichardo-Casas I, Arias C, Vaca L, Zepeda A: Selective distribution and dynamic modulation of miRNAs in the synapse and its possible role in Alzheimer’s Disease. Brain Res. 1584:80–93. 10.1016/j.brainres.2013.12.009

16. Hommers LG, Domschke K, Deckert J: Heterogeneity and Individuality: microRNAs in Mental Disorders. J Neural Transm. 2015, 122:79–97. 10.1007/s00702-014-1338-4

17. Higgins JPT TJCJCMLTPMWV. Cochrane Handbook for Systematic Reviews of Interventions version 6.3 (updated February. (20222022). https://training.cochrane.org/handbook/current.

18. Page MJ, Mckenzie JE, Bossuyt PM, et al.: The PRISMA. 2020. 10.1136/bmj.n71.

19. Cuschieri S: The STROBE guidelines. Saudi J Anaesth. 2019, 13:31. 10.4103/sja.SJA_543_18

20. Hooijmans CR, Rovers MM, Bm De Vries R. Leenaars M, Ritskes-Hoitinga M, Langendam MW. SYRCLE’s risk of bias tool for animal studies [Internet. 2014,

21. Bocchio-Chiavetto L, Maffioletti E, Bettinsoli P, et al.: Blood microRNA changes in depressed patients during antidepressant treatment. European Neuropsychopharmacology. 2013, 23:602–11. 10.1016/j.euroneuro.2012.06.013

22. Homorogan C, Enatescu VR, Nitusca D, et al.: Distribution of microRNAs associated with major depressive disorder among blood compartments. Journal of International Medical Research. 2021, 49:030006052110066. 10.1177/03000605211006633

23. Issler O, Haramati S, Paul ED, et al.: MicroRNA 135 Is Essential for Chronic Stress Resiliency, Antidepressant Efficacy, and Intact Serotonergic Activity. Neuron. 2014, 83:344–60. 10.1016/j.neuron.2014.05.042

24. Enatescu VR, Papava I, Enatescu I, et al.: Circulating Plasma Micro RNAs in Patients with Major Depressive Disorder Treated with Antidepressants: A Pilot Study. Psychiatry Investig. 2016, 13:549. 10.4306/pi.2016.13.5.549

25. Saeedi S, Nagy C, Ibrahim P, et al.: Neuron-derived extracellular vesicles enriched from plasma show altered size and miRNA cargo as a function of antidepressant drug response. Mol Psychiatry. 2021, 26:7417–24. 10.1038/s41380-021-01255-2

26. Fang Y, Qiu Q, Zhang S, et al.: Changes in miRNA-132 and miR-124 levels in non-treated and citalopram-treated patients with depression. J Affect Disord. 2018, 227:745–51. 10.1016/j.jad.2017.11.090

27. Wang X, Wang B, Zhao J, et al.: MiR-155 is involved in major depression disorder and antidepressant treatment via targeting SIRT1. Biosci Rep [Internet. 2018,

28. Kuang WH, Dong ZQ, Tian LT, et al.: MicroRNA-451a, microRNA-34a–5p, and microRNA-221-3p as predictors of response to antidepressant treatment. Brazilian Journal of Medical and Biological Research. 2018, 51:10.1590/1414-431x20187212

29. Lin CC, Tsai MC, Lee C te, et al.: Antidepressant treatment increased serum miR-183 and miR-212 levels in patients with major depressive disorder. Psychiatry Res. 2018, 270:232–7. 10.1016/j.psychres.2018.09.025

30. Fiori LM, Pablo Lopez J, Richard-Devantoy S, et al.: Investigation of miR-1202, miR-135a, and miR-16 in Major Depressive Disorder and Antidepressant Response. International Journal of Neuropsychopharmacology [Internet. 2017, 619-23. 10.1093/ijnp/pyx034

31. Kato M, Ogata H, Tahara H, et al.: Multiple Pre-Treatment miRNAs Levels in Untreated Major Depressive Disorder Patients Predict Early Response to Antidepressants and Interact with Key Pathways. 2022.

32. Kim HK, Tyryshkin K, Elmi N, et al.: Plasma microRNA expression levels and their targeted pathways in patients with major depressive disorder who are responsive to duloxetine treatment. J Psychiatr Res [Internet]. 2019;110: 38–44. Available from: https://linkinghub.elsevier.com/retrieve/pii/S0022395618309646. 10.1016/j.jpsychires.2018.12.007

33. Marshe VS, Islam F, Maciukiewicz M, et al.: Validation study of microRNAs previously associated with antidepressant response in older adults treated for late-life depression with venlafaxine. Prog Neuropsychopharmacol Biol Psychiatry. 2020, 100:109867. 10.1016/j.pnpbp.2020.109867

34. Gururajan A, Naughton ME, Scott KA, et al.: MicroRNAs. 7:862-862. 10.1038/tp.2016.131

35. Belzeaux R, Bergon A, Jeanjean V, et al.: Responder and nonresponder patients exhibit different peripheral transcriptional signatures during major depressive episode. Transl Psychiatry [Internet. 20122185, 10.1038/tp.2012.112

36. Qiao-li Z, Jim L, Xin-yang S, et al.: A preliminary analysis of association between plasma microRNA expression alteration and symptomatology improvement in Major Depressive Disorder (MDD) patients before and after antidepressant treatment. Eur J Psychiatry. 2014, 28:252–64. 10.4321/S0213-61632014000400006

37. He S, Liu X, Jiang K, et al.: Alterations of microRNA-124 expression in peripheral blood mononuclear cells in pre- and post-treatment patients with major depressive disorder. J Psychiatr Res. 2016, 78:65–71. 10.1016/j.jpsychires.2016.03.015

38. Hung YY, Wu MK, Tsai MC, et al.: Aberrant Expression of Intracellular let-7e. 155:647. 10.3390/cells8070647

39. Qi B, Fiori LM, Turecki G, et al.: Machine Learning Analysis of Blood microRNA Data in Major Depression: A Case-Control Study for Biomarker Discovery. International Journal of Neuropsychopharmacology. 202023, 505–10. 10.1093/ijnp/pyaa029

40. Hung YY, Chou CK, Yang YC, et al.: Exosomal let-7e, miR-21-5p, miR-145. 146:2021-9. 10.3390/biomedicines9101428

41. Liu W, Zhang F, Zheng Y, et al.: The role of circulating blood microRNA-374 and microRNA-10 levels in the pathogenesis and therapeutic mechanisms of major depressive disorder. Neurosci Lett. 2021, 763:136184. 10.1016/j.neulet.2021.136184

42. Hong-tao Song, Xin-yang Sun, Wei Niu, et al.: Relationship between the expression level of miRNA-4485 and the severity of depressive symptoms in major depressive disorder patients. The. European Journal of Psychiatry. 2023, 10.1016/j.ejpsy.2022.09.005

43. Ahmadimanesh M, Etemad L, Morshedi Rad D, Ghahremani MH, Mohammadpour AH, Jafarzadeh Esfehani R, Jowsey P, Behdani F, Moallem SA, Abbaszadegan MR. Administration of melatonin protects against acetylsalicylic acid-induced impairment of male reproductive function in mice. Iran J Basic Med Sci 2023; 26: 820–829. doi: 10.22038/IJBMS.2023.66496.14595

44. Wang Y, Yan Y, Wei J, et al. Down-regulated miR-16-2 in peripheral blood is positively correlated with decreased bilateral insula volume in patients with major depressive disorder. J Affect Disord. 2023;338:137–143. doi:10.1016/j.jad.2023.05.068

45. Funatsuki T, Ogata H, Tahara H, Shimamoto A, Takekita Y, Koshikawa Y, Nonen S, Higasa K, Kinoshita T, Kato M. Changes in Multiple microRNA Levels with Antidepressant Treatment Are Associated with Remission and Interact with Key Pathways: A Comprehensive microRNA Analysis. Int J Mol Sci. 2023 Jul 30;24(15):12199. doi: 10.3390/ijms241512199. PMID: 37569574; PMCID: PMC10418406.

46. Ogata H, Higasa K, Kageyama Y, et al. Relationship between circulating mitochondrial DNA and microRNA in patients with major depression. J Affect Disord. 2023;339:538–546. doi:10.1016/j.jad.2023.07.073

47. Patrício P, Mateus-Pinheiro A, Alves ND, et al.: miR-409 and miR-411 Modulation in the Adult Brain of a Rat Model of Depression and After Fluoxetine Treatment. Front Behav Neurosci. 2020, 14:10.3389/fnbeh.2020.00136

48. lo Iacono L, Ielpo D, Parisi C, et al.: MicroRNA-34a regulates 5-HT2C expression in dorsal raphe and contributes to the anti-depressant-like effect of fluoxetine. Neuropharmacology. 2021, 190:108559. 10.1016/j.neuropharm.2021.108559

49. Zhang Y, Wang Y, Wang L, et al.: Dopamine Receptor D2 and Associated microRNAs Are Involved in Stress Susceptibility and Resistance to Escitalopram Treatment. International Journal of Neuropsychopharmacology [Internet. 2015110,

50. Song W, Shen Y, Zhang Y, et al.: Expression alteration of microRNAs in Nucleus Accumbens is associated with chronic stress and antidepressant treatment in rats. 2019.

51. Xie L, Chen J, Ding YM, et al.: MicroRNA-26a-2 maintains stress resiliency and antidepressant efficacy by targeting the serotonergic autoreceptor HTR1A. Biochem Biophys Res Commun. 2019, 511:440–6. 10.1016/j.bbrc.2019.02.078

52. Grieco SF, Velmeshev D, Magistri M, et al.: Ketamine up-regulates a cluster of intronic miRNAs within the serotonin receptor 2C gene by inhibiting glycogen synthase kinase-3. The World Journal of Biological Psychiatry. 2017, 18:445–56. 10.1080/15622975.2016.1224927

53. Wan YQ, Feng JG, Li M, et al.: Prefrontal cortex miR-29b-3p plays a key role in the antidepressant-like effect of ketamine in rats. Exp Mol Med. 2018, 50:1–14. 10.1038/s12276-018-0164-4

54. Mingardi J, la Via L, Tornese P, et al.: miR-9-5p is involved in the rescue of stress-dependent dendritic shortening of hippocampal pyramidal neurons induced by acute antidepressant treatment with ketamine. Neurobiol Stress. 2021, 15:100381. 10.1016/j.ynstr.2021.100381

55. Huang C, Wang Y, Wu Z, et al.: miR-98-5p plays a critical role in depression and antidepressant effect of ketamine. Transl Psychiatry. 2021, 11:454. 10.1038/s41398-021-01588-0

56. Pan B, Liu Y: Original Article Effects of duloxetine on microRNA expression profile in frontal lobe and hippocampus in a mouse model of depression [Internet]. Vol. 8. Int J Clin Exp Pathol. 2015,

57. Higuchi F, Uchida S, Yamagata H, et al.: Neurobiology of Disease Hippocampal MicroRNA-124 Enhances Chronic Stress Resilience in Mice. 2016. 10.1523/JNEUROSCI.0319-16.2016

58. Zeng D, Shi Y, Li S, et al. miR-124 Exacerbates depressive-like behavior by targeting Ezh2 to induce autophagy. Behav Pharmacol. 2023;34(2-3):131–140. doi:10.1097/FBP.0000000000000716

59. Guan W, Wu XY, Jin X, Sheng XM, Fan Y. miR-204–5p Plays a Critical Role in the Pathogenesis of Depression and Anti-depression Action of Venlafaxine in the Hippocampus of Mice [published online ahead of print, 2023 Jun 23]. Curr Med Chem. 2023;10.2174/0929867330666230623163315. doi:10.2174/0929867330666230623163315

60. Oved K, Morag A, Pasmanik-Chor M, et al.: Genome-wide miRNA expression profiling of human lymphoblastoid cell lines identifies tentative SSRI antidepressant response biomarkers. Pharmacogenomics. 2012, 13:1129–39. 10.2217/pgs.12.93

61. Oved K, Morag A, Pasmanik-Chor M, et al.: Genome-wide expression profiling of human lymphoblastoid cell lines implicates integrin beta-3 in the mode of action of antidepressants. Transl Psychiatry [Internet. 20133, 10.1038/tp.2013.86

62. Angelucci F, Croce N, Spalletta G, et al.: Paroxetine Rapidly Modulates the Expression of Brain-Derived Neurotrophic Factor mRNA and Protein in a Human Glioblastoma-Astrocytoma Cell Line. Pharmacology. 2011:5–10. 10.1159/000322528

63. Wibrand K, Pai B, Siripornmongcolchai T, et al.: MicroRNA regulation of the synaptic plasticity-related gene Arc. PLoS One. 2012, 7:10.1371/journal.pone.0041688

64. Sonntag KC, Woo TUW, Krichevsky AM, et al.: Converging miRNA functions in diverse brain disorders: A case for miR-124 and miR-126. Exp Neurol. 2012, 235:427–35. 10.1016/j.expneurol.2011.11.035

65. Chandrasekar V, Dreyer JL: microRNAs miR-124, let-7d and miR-181a regulate Cocaine-induced Plasticity. Molecular and Cellular Neuroscience. 2009, 42:350–62. 10.1016/j.mcn.2009.08.009

66. Shi LS, Ji CH, Tang WQ, et al.: Hippocampal miR-124 Participates in the Pathogenesis of Depression via Regulating the Expression of BDNF in a Chronic Social Defeat Stress Model of Depression. Curr Neurovasc Res. 2022, 19:210–218. 10.2174/1567202619666220713105306

67. Anaïs A, Arnould T, Renard P: SURVEY AND SUMMARY miR-212/132 expression and functions: within and beyond the neuronal compartment. Available from.

68. Lopez JP, Lim R, Cruceanu C, et al.: 1202, 2014:764–8. 10.1038/nm.3582

69. Penner-Goeke S, Binder EB: Dialogues in Clinical Neuroscience Epigenetics and depression Epigenetics and depression. DIALOGUES IN CLINICAL NEUROSCIENCE • [Internet. 201921, 397–405.

70. Choudrie J, Dwivedi YK: Investigating the research approaches for examining technology adoption issues [Internet]. Vol. 1, Journal of Research Practice. 200547, 4:7.

